# Directional anchor genes refine polygenic informed treatment selection in schizophrenia and bipolar disorder

**DOI:** 10.1101/2022.03.20.22272666

**Authors:** William R. Reay, Michael P. Geaghan, Joshua R. Atkins, Vaughan J. Carr, Melissa J. Green, Murray J. Cairns

## Abstract

Genetically informed drug development and repurposing is an attractive prospect for improving outcomes in patients with psychiatric illness; however, the effectiveness of these endeavours can be confounded by heterogeneity. In this study, we propose a novel approach that links interventions implicated by disorder-associated genetic risk, at the population level, to a framework that can target these compounds to individuals. Specifically, results from genome-wide association studies are integrated with expression data to prioritise individual “directional anchor” genes for which the predicted risk-increasing direction of expression could be counteracted by an existing drug. While these compounds represent plausible therapeutic candidates, they are not likely to be equally efficacious for all individuals. To account for this heterogeneity, we constructed polygenic scores restricted to variants annotated to the network of genes that interact with each directional anchor gene. These metrics, we call pharmagenic enrichment scores (PES), identify individuals with a higher burden of genetic risk, localised in biological processes related to the candidate drug target, to inform precision drug repurposing. We used this approach to investigate schizophrenia and bipolar disorder and reveal several compounds targeting specific directional anchor genes that could be plausibly repurposed, including antioxidants, vitamins, antiarrhythmics, and lipid modifying agents. These genetic risk scores, mapped to the networks associated with target genes, revealed biological insights that cannot be observed in undifferentiated genome-wide polygenic risk score (PRS). For example, an enrichment of these partitioned scores in schizophrenia cases with otherwise low PRS and distinct correlations with measured biochemical traits. In summary, genetic risk could be used more specifically to direct drug repurposing candidates that target particular genes implicated in psychiatric and other complex disorders.

## INTRODUCTION

Psychiatric disorders remain difficult to effectively manage in some patients, with treatment resistance observed in a notable proportion of individuals prescribed conventional pharmacotherapies [1–3]. Moreover, a key challenge in psychiatric practice is the selection of a suitable course of treatment for newly diagnosed patients. Novel treatment opportunities for these disorders would be of great clinical benefit, however, the drug development pipeline remains arduous, expensive, and unproductive [4, 5]. Drug repurposing, whereby an approved compound is redeployed for a new indication, is a promising avenue to more rapidly alter psychiatric practice relative to the *de novo* drug development process [6, 7]. There has already been utility in this approach demonstrated in psychiatry, such as atomoxetine that has been repurposed for attention-hyperactive deficit disorder (ADHD) and the anti-convulsant valproate for bipolar disorder [8].

We know that psychiatric illnesses arise from a multifaceted interplay between genetic and environmental factors that contribute to its aetiologic complexity. In recent years, genome-wide association studies (GWAS) have confirmed that psychiatric disorders are polygenic in nature [9–13], with common frequency variants constituting a significant portion of trait heritability. This means that individual loci that have small to modest impact, but contribute to a much larger polygenic effect of many such variants throughout the genome [14, 15]. Biological insights from genetic studies could lead to repurposing opportunities in psychiatry – for example, schizophrenia GWAS have previously suggested that the common variant signal is enriched amongst the targets of antiepileptics, as well as in genes involved in retinoid (vitamin A derivative) pathways [16, 17]. The polygenic nature of these disorders, however, present a challenge drug targeting because the genetic architecture of each individual will be highly heterogeneous. This means that any given patient will carry a unique combination of risk and protective alleles, which likely translates to different underlying biological processes being affected. As a result, genetically informed drug candidates may not be efficacious at a population level. These phenomena necessitate the consideration of how pharmacotherapies could be targeted more specifically to individuals based on their underlying genetic and clinical risk factors.

Our group has previously sought to address these challenges through the development of the *pharmagenic enrichment score* (PES), which is a framework that seeks to use polygenic risk to direct precision drug repurposing opportunities [7, 18–20]. Specifically, the PES approach derives partitioned polygenic scores from variants annotated to pathways or networks that are targeted by approved drugs, with the underlying hypothesis that individuals with elevated genetic risk (PES) amongst those genes may benefit from a compound which modulates that pathway. Furthermore, in prior work we have established that PES profiles provide distinct insights from a biologically undifferentiated genome-wide polygenic risk score (PRS) [18–20]. However, a limitation of the PES approach is that it is not innately informative as to which of the suite of drugs targeting a pathway will be most useful, particularly in regards as to whether an agonist or antagonist of target genes should be investigated. While we addressed this previously, in respiratory medicine, by triangulating prioritised pathways through causal inference of pharmacologically sensitive biochemical traits [19, 20], these relationships are more difficult to find in psychiatric disorders [21]. In the current study, we propose a new implementation of the PES that is informed by genetically proxied mRNA or protein expression of drug target genes. This approach identifies candidate psychiatric drug repurposing opportunities at the population level that can then be more appropriately integrated with genetic risk scores relevant to these target genes, to identify individuals who may benefit more readily from these compounds. These are termed *directional anchor genes* as they inform on the clinically useful direction of modulation is for the biological networks containing these genes which can be utilised to construct PES. In this study, we investigated this novel approach in two highly heritable psychiatric disorders, schizophrenia and bipolar disorder, and identified novel drug repurposing opportunities from candidate directional anchor genes and propose how these genes could be used in concert with the PES to direct the candidate compounds for repurposing.

## MATERIALS AND METHODS

### Overview of the directional anchor gene *pharmagenic enrichment score* approach

We summarise the fundamental principles of directional anchor genes and their integration with the *pharmagenic enrichment score* in this section, followed by more expansive details in the subsequent sections. Briefly, we define the concept of a directional anchor gene (DA-gene) as a gene where, i) the direction of expression associated with increased odds of the disorder can be predicted, and ii) this disorder associated direction of expression could be counteracted by an approved compound, thus constituting a drug repurposing opportunity. For instance, if upregulation of a hypothetical gene, gene *X*, was associated with greater odds of a disease phenotype, then an antagonist of gene *X* may be clinically useful. If this gene *X* antagonist is already approved for another indication, this may inform drug repurposing. However, there is immense heterogeneity between individuals for any given complex trait or disease in its genetic architecture, which often translates to highly variable clinical manifestation. We therefore hypothesise that individuals with a greater burden of disorder-associated polygenic risk in the directional anchor gene, and its network of genes that physically and biologically interact with it, may benefit more specifically from a drug repurposing candidate targeting the DA-gene. Polygenic risk mapped to biological networks encompassing the directional anchor genes is likely to incorporate disorder-associated impacts on upstream processes that would modify the effect of a compound targeting the candidate gene, as well as downstream processes triggered by modulating the directional anchor. As discussed in the introduction, our group has previously developed the *pharmagenic enrichment score* (PES) methodology to utilise polygenic scoring to direct drug repurposing, whereby polygenic scores are constructed specifically using variants mapped to biological pathways targeted by known drugs [18, 19]. A limitation of the PES approach is that it does not inherently predict the direction of effect genes in the pathway that would need to be targeted such that repurposing a drug for individuals with high polygenic risk in said pathway would be efficacious. Directional anchor genes, therefore, help address this limitation when used in conjunction with PES constructed using networks or biological pathways in which the gene participates. In other words, drug repurposing opportunities are predicted at the population level based on the expression of target genes, with these compounds potentially able to be more specifically directed to individuals with elevated disorder associated genetic risk within pathways or networks that contain the directional anchor gene, that is, an elevated PES (Figure 1). It should be noted that whilst we apply this approach to binary disease phenotypes in this study, it can also be utilised for clinically relevant continuous traits. In that case, candidate directional anchor genes would be those genes for which the drug repurposing candidate is genetically inferred to modulate the trait in a clinically useful fashion.

**Figure 1.**
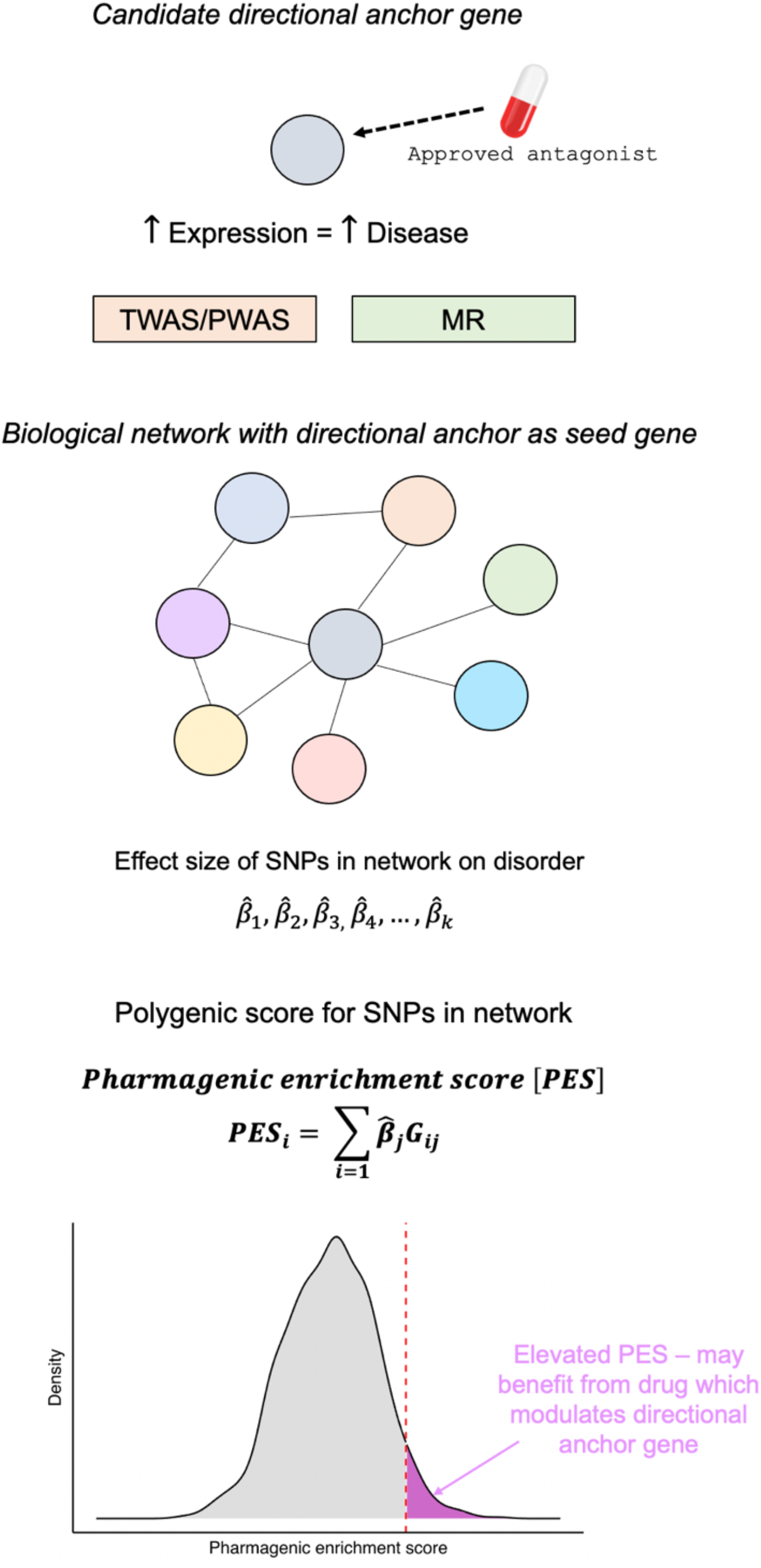
Overview schematic of the integration of candidate directional anchor genes with *pharmagenic enrichment scores*. Directional anchor genes are genes targeted by an approved compound in what is genetically predicted through integration with expression data to decrease the risk of the disorder or modulate the trait, if continuous, in a clinically useful manner. For instance, if increased expression of a gene is associated with a disorder through a transcriptome or proteome-wide association study (TWAS/PWAS) or Mendelian randomisation (MR) using quantitative trait loci as instrumental variables, then an antagonist of said gene may be a repurposing opportunity. Directional anchor genes then act as seed genes to define a network of other genes that interact with them. SNPs mapped to this network are then utilised to construct a *pharmagenic enrichment score* (PES) for the network. In the case of a binary disease phenotype, the interpretation of the PES would be that individuals with an elevated score relative to an appropriate population reference may benefit from a compound which modulates the directional anchor gene. The hypothesis underlying this is that these individuals will have genetic risk that impacts upstream or downstream processes relating to the directional anchor, as well as the anchor gene itself, which could be addressed by the repurposing candidate in question.

### Schizophrenia and bipolar disorder genome-wide association studies

We obtained GWAS summary statistics for schizophrenia (SZ) and bipolar disorder (BIP) from the psychiatric genomics consortium [9, 10]. The SZ GWAS was a mega-analysis of mostly European ancestry cohorts and comprised 67,390 cases and 94,015 controls, whilst the European ancestry BIP GWAS mega-analysis had 20,352 cases and 31,358 controls. In addition, we also utilised the same SZ GWAS with a constituent cohort removed (Australian Schizophrenia Research Bank) when we profiled PES within that dataset, as described in a proceeding section of the methods.

### Transcriptome and proteome-wide association studies

A transcriptome-wide association study (TWAS) and a proteome-wide association study (PWAS) was performed of SZ and BIP by leveraging genetically imputed models of mRNA and protein expression, respectively. Specifically, we utilised the FUSION approach for TWAS/PWAS, with full details outlined in the supplementary materials [22]. Expression weights for the TWAS were derived from postmortem brain (GTEx v7, PsychENCODE) and whole blood (GTEx v7), whilst protein expression weights were similarly from postmortem brain (ROSMAP) and whole blood (ARIC) [22–25]. The FUSION methodology integrates SNP-effects from the model of genetically predicted expression with the effects of the same SNPs on SZ or BIP, after accounting for linkage disequilibrium, such that the TWAS *Z* score can be a conceptualised measure of genetic covariance between mRNA or protein expression of the gene and the GWAS trait of interest. We utilised a conservative method for multiple-testing correction whereby the Bonferroni methodology was implemented to divide the alpha level (0.05) by the total number of significantly *cis*-heritable models of genetically regulated expression (GReX) tested from any brain tissue considered or whole blood (Supplementary Text, Supplementary Tables 1-4). Several genes had GReX available in multiple-tissues, thus rendering Bonferroni correction conservative, however, we implemented this approach to capture only the most confidently associated genes that could constitute drug-repurposing candidates. For candidate directional anchor genes derived from TWAS/PWAS, we probabilistically finemapped those regions using the FOCUS methodology using the default prior (*p* = 1 × 10^−3^) and prior variance (*nσ*^2^ = 40) to approximate Bayes’ factors, such that the posterior inclusion probability (*PIP*) of each gene being a member of a credible set with 90% probability of containing the causal gene could be derived [26]. We also investigated the impact of using a more conservative prior as outlined in the supplementary text. Moreover, we tested whether SNPs that constitute the GReX model and either SZ or BIP displayed statistical colocalisation with the *coloc* package as implemented by FUSION [27].

### Mendelian randomisation

In addition, we leveraged variants strongly correlated with mRNA (expression quantitative trait loci – eQTL) and protein expression (protein expression quantitative trait loci – pQTL), respectively, as instrumental variables (IVs) in a two-sample Mendelian randomisation (MR) analysis [28]. Analogous to the TWAS/PWAS, eQTL/pQTL were derived from brain (MetaBrain, ROSMAP) and blood (eQTLGen, Zhang *et al*.), with full details described in the supplementary text [28–31]. Strict selection criteria were implemented to select suitable IVs, including only retaining independent genome-wide significant (*P* < 5 × 10^−8^) SNPs that were associated with three or less mRNA/proteins in each relevant tissue/study (Supplementary Text). Moreover, we utilised a more stringent LD clumping procedure for eQTLs given the greater power and sample sizes for these datasets also results in immense pleiotropy amongst the SNP effects on mRNA. This was achieved by only selecting the most significant independent SNPs using one megabase clumps, with LD estimated using the 1000 genomes phase 3 panel. The effect of mRNA or protein expression for any given gene on SZ or BIP was estimated using the Wald ratio (single IV) or an inverse-variance weighted estimator (multiple IVs, with fixed effects due to the small number of IVs). As in the TWAS/PWAS, we utilised Bonferroni correction across all tissues in the mRNA and protein analyses respectively and then sought to identify candidate directional anchor genes from these signals. For any candidate directional anchor genes, where an approved drug was predicted to reverse the odds increasing direction of expression, we performed a series of sensitivity analyses (Supplementary Text). Briefly, these involved: assessing the genomic locus of the IV SNP, for genes it may be associated with, by testing for evidence of a shared causal variant through colocalisation (default priors) and conducting a phenome-wide Mendelian randomisation analysis (MR-pheWAS) using SNP effects from each trait in the IEUGWAS database. The above MR and sensitivity analyses were performed using the R packages TwoSampleMR v0.5.5, ieugwasr v0.1.5, and coloc v4.0.4 [27, 32].

### Identifying drug repurposing candidates

We searched genes prioritised from the TWAS/PWAS or MR analyses in the Drug-gene interaction database (DGIdb v4.2.0 – accessed April 2021) to identify approved compounds that could reverse the odds increasing direction of expression for SZ or BIP [33]. DGIdb combines data from databases such as DrugBank, as well as curated literature sources. We defined high confidence drug-gene interactions as those reported in DrugBank as well as at least one other database or literature source.

### Identification of genes interacting with candidate directional anchor genes

Protein-protein interaction data was downloaded from the STRING database v11 [34]. We utilised each of the six candidate directional anchor genes as a seed gene, separately, and constructed a network of genes predicted to interact with the seed gene by retaining high confidence edges (confidence score > 0.7) derived from experimental evidence or curated protein-complex and pathway databases, as this is generally considered the most rigorous evidence from STRING. We then tested which gene-sets curated by the g:Profiler (version e104_eg51_p15_3922dba) resource (GO, KEGG, Reactome, WikiPathways, TRANSFAC, miRTarBase, Human Protein Atlas, CORUM, and Human phenotype ontology) were overrepresented amongst the genes in each network, using the g:SCS (set counts and sizes) multiple-testing correction method implemented by g:Profiler that has been shown to better account for the complex, overlapping nature of these data [35]. We considered a corrected *P* value < 0.05 as significant.

We then tested the association of the genes in each of these networks, with and without the gene removed, with the common variant signal in the SZ and BIP GWAS using MAGMA v1.09 [36]. Briefly, SNP-wise *P* values were aggregated at gene-level, with SNPs annotated to genes using two different sets of genic boundary extensions to capture potential regulatory variation, conservative (5 kilobases (kb) upstream, 1.5 kb downstream), and liberal (35 kb upstream, 10 kb downstream). Gene-set association is implemented by MAGMA using linear regression, whereby the probit transformed genic *P* values (*Z* scores) are the outcome with a binary explanatory variable indicating whether a gene is in the set to be tested (*β*_*S*_), covaried for other confounders like gene size, as described previously. The test statistic of interest is a one-sided test of whether *β*_*S*_ > 0, and thus, quantifies if the genes in the set are more associated than all other genes. We also investigated the association of the approximately 34,000 gene-sets collated by g:Profiler, such that we could demonstrate whether gene-sets overrepresented in each network were also associated with SZ or BIP.

### Constructing directional anchor gene network PES

We sought to utilise variants annotated to the genes within the network of each candidate directional anchor genes to develop *pharmagenic enrichment scores* for SZ and BIP, respectively. As described previously, a PES is analogous to a genome-wide PRS in its derivation, with the key defence that it only utilises variants mapped to the gene-set of interest (equation 1) [18]. Specifically, a PES profile in individual *i* comprises the sum of the effect size of *j* variants from the GWAS 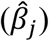 annotated to at least one gene in set *M*, multiplied by the allelic dosage under an additive model (*G*_*ij*_ *∈ G* = 0, 1, 2).

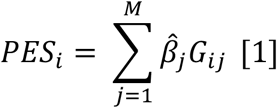

The genome wide PRS for SZ and BIP are essentially the same model but *M* incorporates the entire genome. In accordance with the MAGMA analyses, we tested two genic boundary configurations for evaluating the best performing PES for each directional anchor gene network – conservative and liberal. Our previous PES related approaches utilised the LD clumping and thresholding (C+T) approach, whereby SNPs are ‘clumped’ such that the retained SNPs are largely independent and ‘thresholded’ based on their association *P*-value in the GWAS. In each case the threshold was set at the optimum for the druggable gene-set association at the population level. However, given we selected the gene-sets in this study based on interactions with the candidate directional anchor gene, we tested four different *P* value thresholds (*P*_*T*_ *∈ T* = {0.005, 0.05, 0.5, 1}), which represent a model with all SNPs, nominally significant SNPs, and a threshold an order of magnitude above or below the nominal threshold. These choices of *T* have been discussed extensively elsewhere [18–20]. We utilised PRSice-2 v2.3.3 (linux) for the C+T models [37]. In addition, we utilised a penalised regression framework to shrink SNP effect sizes to optimise the model for each PES, as implemented by the standalone version of lassosum v0.4.5 [38]. The implementation for this method has been outlined extensively elsewhere, with the optimal tuning parameter (*λ*) based on the score that displays the highest correlation with the phenotype and the best performing constraint parameter (*s*) chosen from a range of *a priori* specified values to decrease computational burden (0.2, 0.5, 0.9, and 1).

### Training and validation of directional anchor gene network PES

We utilised the prospective UK Biobank (UKBB) cohort to define the best performing PES for each directional anchor gene network. Our group has previously processed the UKBB genotype data such that unrelated individuals of white British ancestry were retained, along with other sample and variant level quality control considerations applied [20]. As a result, the composition of the full UKBB cohort in this study was 336,896 participants for which up to 13,568,914 autosomal variants were available (imputation INFO > 0.8). SZ and BIP cases were defined in the UKBB using a combination of self-report data both from the general assessment visit and the mental health questionnaire (MHQ), along with hospital inpatient data (primary or secondary ICD-10 codes), with full details in the supplementary text. In total, there were 631 UKBB participants from the study cohort defined as having SZ, with 1657 BIP cases identified. The controls were double the number of the respective case cohorts randomly, and independently for SZ and BIP, derived from 75,201 individuals with genotype data that completed the MHQ and did not self-report any mental illness. The full complement of SZ cases with the aforementioned controls (N = 1262) was utilised as the training set for the SZ scores given the relatively small number of cases. As a result, we utilised the Australian Schizophrenia Research Bank (ASRB) cohort as a validation set to attempt to replicate the associations observed with the scores, which has been described elsewhere [18, 39]. The ASRB was a component of the PGC3 SZ GWAS, and thus, we retrained all the best performing PES scores using summary statistics with the ASRB cohort removed before testing them in that dataset. The BIP analyses employed a 70/30 split for the training and validation cohort in the UKBB, with double the number of independent MHQ derived healthy controls utilised for each case-set. Further information regarding the demographic composition of these cohorts is provided in the supplementary text.

The PES and PRS constructed using the C+T configurations and penalised regression were scaled to have a mean of zero and unit variance before evaluating their association with SZ or BIP, for the respective scores in the UKBB training cohorts, using binomial logistic regression covaried for sex, age, genotyping batch, and five SNP derived principal components. The optimal PES for each network was selected for each disorder separately by calculating the variance explained on the liability scale (Nagelkerke’s *R*^2^), assuming a 0.7% and 1% prevalence for SZ and BIP, respectively [40]. These PES/PRS that explained the most phenotypic variance were then profiled and tested in the validation sets. For PES that were significantly associated with either disorder, we conservatively constructed another model that also included genome wide PRS, with a *χ*^2^ test of residual deviance performed to ascertain whether adding the PES in addition to the PRS significantly improved model fit. Correlations (Pearson’s) amongst scaled PES and PRS were visualised using the corrplot package v0.84. Individuals with at least one elevated PES in the training cohorts (highest decile) were identified, with this binary variable tested for association with SZ or BIP using another logistic regression model. Finally, we also considered *residualised* PES, whereby the residuals were extracted and scaled from a linear model that regressed genome wide PRS against principal components and genotyping batch on the score in question. All analyses described in this paragraph were performed utilising R v3.6.0.

### Biochemical and mental health phenome-wide association studies

We then investigated the correlations between the best performing PES for each network and i) blood or urine biochemical traits, and ii) self-reported mental health disorders besides SZ or BIP. The biochemical analyses were performed in up to 70,625 individuals who did not self-report any mental health disorders in the MHQ and were also not included in the SZ or BIP training/validation sets as controls. There were 33 biochemical traits tested (raw values - Supplementary Table 5) in a linear model with each PES or PRS as an explanatory variable covaried for sex, age, sex x age, age^2^, 10 principal components, and genotyping batch. We also performed sex-stratified analyses, with oestradiol additionally considered in females. A number of sensitivity analyses were performed for PES-biochemical trait pairs that were significantly correlated after FDR correction– i) adjustment for genome-wide PRS, ii) natural log transformation of the biochemical outcome variable, iii) inverse-rank normal transformed residuals as the outcome variable from a model that regressed sex, sex x age, and age^2^, and iv) adjustment for statin use (given the number of lipid related signals uncovered). These correlations are observational in nature, and thus, there are several other potential confounders that could be considered – however, given the potential biases induced by adjusting for heritable covariates, we utilised the above strategies as a baseline suite of sensitivity analyses [41]. A specific test of sexual dimorphisms between the regression results in males and females was also performed based on the sex-specific regression estimates and standard errors, as outlined elsewhere [42]. Moreover, we then evaluated the association between each score and 14 non-SZ or BIP mental health disorders which individuals who completed the MHQ were asked to self-report (Supplementary Table 6). In all instances, we used the 70,625 individuals who did not self-report any mental disorders as the controls in binomial logistic regression models covaried for the same terms as in the biochemical analyses.

## RESULTS

### Candidate directional anchor genes reveal novel drug repurposing opportunities in psychiatry

We sought to identify candidate directional anchor genes for SZ or BIP by integrating GWAS summary statistics for these traits with transcriptomic and proteomic data collected from either blood or post-mortem brain (Figure 2a). Specifically, we utilised genetically imputed models of mRNA or protein expression to conduct a TWAS and PWAS, respectively (Supplementary Tables 1-4). Genome-wide significant eQTLs and pQTLs were also leveraged as instrumental variables in a more conservative two-sample Mendelian randomisation analysis to explicitly test for any causal effects of mRNA or protein expression (Supplementary Tables 7-10). After implementing Bonferroni correction within each analysis set (TWAS, PWAS, eQTL-MR, pQTL-MR), we found several genes for which their expression was associated with at least one of the psychiatric phenotypes at the mRNA or protein level that was also putatively modulated by an approved compound in a risk decreasing direction. There were 13 druggable genes from TWAS for which the direction of genetically predicted mRNA expression correlated with SZ could be pharmacologically counteracted, whilst there were two such genes for BIP, some examples of which are visualised in Figure 2b. For instance, imputed mRNA expression of the calcium voltage-gated channel subunit gene *CACNA1C* was negatively correlated with SZ (*P* = 3.65 × 10^−15^), and thus, an activator of this gene like the antiarrhythmic agent Ibutilide may be a repurposing candidate for SZ, with this gene also trending towards association with BIP in the same direction (*P* = 3.18 × 10^−5^). We compared the TWAS results to that of a PWAS using data from blood or brain tissue, although the number of proteins assayed in these studies was considerably smaller than that of the number of mRNA available, and thus, most of the candidate genes derived using TWAS did not have protein measurements available for a direct comparison of the effect of protein expression relative to mRNA. However, there were two Bonferroni significant TWAS genes that represented a plausible repurposing candidate with protein expression data available (*NEK4* and *CTSS*), with both genes showing a similar strength of association in the PWAS. For any given genetically predicted mRNA or protein expression, the gene is not necessarily causal due to LD complexity and other phenomena such as co-regulation [43]. This is critical for using these approaches to direct drug repurposing as the target the genes need to correspond to the genetic association for the disorder. As a result, we implemented a Bayesian finemapping procedure for each TWAS candidate gene to identify plausible causal genes in each locus (Supplementary Text). We found four repurposing candidate genes for SZ with strong evidence for membership of a credible set with 90% probability of containing the causal gene (*PIP* > 0.8 – *PCCB, GRIN2A, FES*, and *CACNA1D*). However, *CACNA1D* was excluded from further analyses due to the poor performance of its imputed model and complexity of its locus on chromosome three, as outlined more extensively in the supplementary text. We then considered a more lenient posterior inclusion probability of 0.4, which identified two more genes for SZ (*CACNA1C* and *RPS17*), and a BIP gene (*FADS1*). Colocalisation analyses were also performed to test a related, but distinct hypothesis that the GWAS signal and SNP weights in the expression model share an underlying single causal variant. Interestingly, for the genes selected using the lower confidence *PIP* > 0.4 threshold, we found strong evidence for a shared causal variant (*PP*_H4_ > 0.9), supporting their inclusion as putative drug repurposing targets. We did not consider the two genes shared with the PWAS any further as they did not display strong finemapping support in the TWAS, which is a more accurate representation of any given locus due to the more expansive number of genes with RNAseq available. In summary, using a genetically imputed expression approach we identified five candidate directional anchor genes for SZ and one for BIP (Table 1). For example, imputed *GRIN2A* mRNA expression was negatively correlated with SZ (*P* = 1.44 × 10^−9^), with a trend also observed for BIP (*P* = 5.07 × 10^−3^), with compounds of interest in psychiatry, such as *N*-acetylcysteine, known to agonise this subunit [44, 45].

**Table 1.** Candidate directional anchor genes for schizophrenia and bipolar along with their associated drug repurposing candidates.

| Gene | Disorder | Protective direction | Repurposing candidates | Sensitivity analyses |
| --- | --- | --- | --- | --- |
| *PCCB* | SZ | Increased expression | Biotin | High confidence causal gene from TWAS ($PIP > 0.8$). Further supported by MR |
| *FADS1* | BIP | Increased expression | Icosapent<br>Linolenic acid | Moderate confidence causal gene from TWAS ($PIP > 0.4$, additionally supported by colocalisation – $PP_{H4} > 0.8$). Further supported by MR |
| *GRIN2A* | SZ | Increased expression | *N*-acetylcysteine<br>Glycine | High confidence causal gene from TWAS ($PIP > 0.8$). |
| *CACNA1C* | SZ | Increased expression | Ibutilide | Moderate confidence causal gene from TWAS ($PIP > 0.4$, additionally supported by colocalisation – $PP_{H4} > 0.8$). |
| *RPS17* | SZ | Increased expression | Artenimol | Moderate confidence causal gene from TWAS ($PIP > 0.4$, additionally supported by colocalisation – $PP_{H4} > 0.8$). |
| *FES* | SZ | Decreased expression | Fostamatinib<br>Lorlatinib | High confidence causal gene from TWAS ($PIP > 0.8$). |

**Figure 2.**
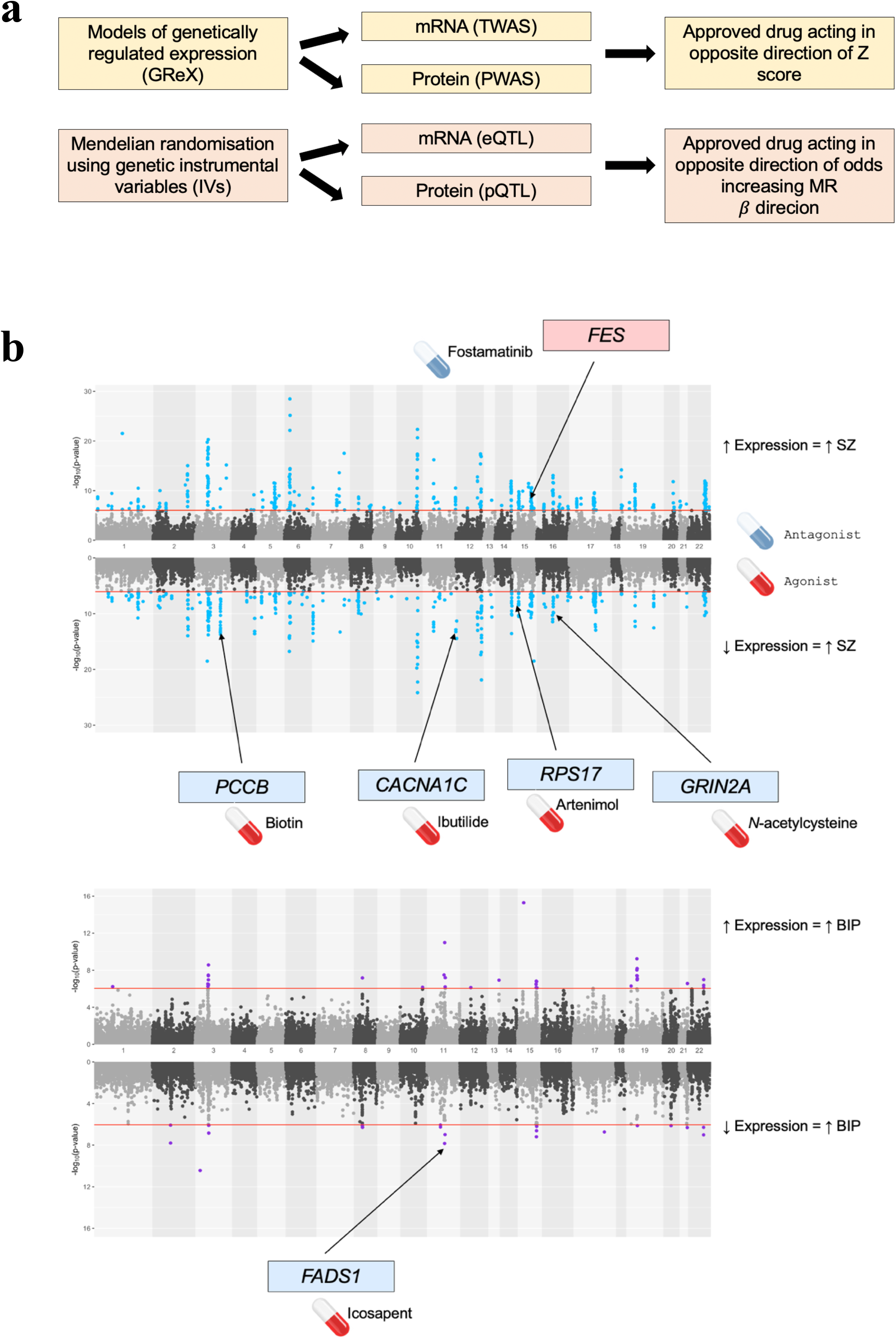
Identification of candidate directional anchor genes for schizophrenia and bipolar through integration of transcriptomic and proteomic data. (**a**) Schematic for the prioritisation of candidate directional anchor genes through models of genetically regulated expression (GReX, yellow) and Mendelian randomisation (orange). In both instances, approved compounds are derived for implicated genes that reverse the odds increasing direction of mRNA or protein expression. TWAS = transcriptome-wide association study, PWAS = proteome-wide association study (**b**) Results of the multi-tissue (brain and blood) TWAS for schizophrenia (SZ, top) and bipolar disorder (BIP, bottom). The Miami plot visualises the -log10 transformed *P* value of association with genes exhibiting a negative genetic covariance between expression and the trait, that is, TWAS *Z* < 0, plotted in the downward direction. The red line denotes the Bonferroni threshold. The candidate directional anchor genes from the TWAS approach are highlighted on the plot along with their putative repurposing candidate that corrects the odds-increasing direction of expression. For example, predicted *PCCB* expression is negatively correlated with SZ, and thus, a *PCCB* agonist like biotin may be clinically useful.

To prioritise candidate directional anchor genes, we utilised eQTL and pQTL as IVs to estimate the causal effect of mRNA or protein expression on either disorder outcome in a Mendelian randomisation analysis, given more onerous assumptions are met (Supplementary Materials, Supplementary Tables 7-10). This is critical as the use of molecular QTLs related to variables like mRNA expression as IVs is challenging due to LD complexity and the potential effect of QTLs on multiple genes [28, 46]. As a result, we sought to complement the above discovery orientated TWAS/PWAS with more conservative selection criteria for an eQTL or pQTL to be an IV, particularly in the case of eQTLs where sample sizes for some tissues are now very large. Independent SNPs (LD *r*^2^ < 0.001) acting as eQTLs or pQTLs at a threshold of genome-wide significance (*P* < 5 × 10^−8^) were selected from post-mortem brain or blood, as outlined in the methods and supplementary materials. Due to the conservative nature of these analyses, many of the genes considered in the TWAS/PWAS did not have a suitable IV available. Conversely, a small number of genes that did not display adequate multivariate *cis*-heritability in the TWAS/PWAS weights could now be included. The mRNA models after Bonferroni correction uncovered four genes for which expression exerted a potential causal effect on SZ or BIP with a suitable compound approved to reverse the odds-increasing direction of effect. There were three for SZ (*PCCB, NEK1*, and *PTK2B*), as well as *FADS1* for bipolar. Interestingly, *PCCB* and *FADS1* overlapped with the TWAS results – as an example, each standard deviation increase in cortical *FADS1* expression was associated with approximately 15.23% [95% CI: 8.69%, 21.77%] decrease in the odds of BIP, which could be accentuated by a *FADS1* agonist like the omega-3 fatty acid supplement icosapent (Ethyl eicosapentaenoic acid). We then performed a series of sensitivity analyses to assess IV validity and for evidence of confounding pleiotropy (Supplementary Materials). These analyses supported *PCCB* and *FADS1* as candidate directional anchor genes. The index IV-SNPs mapped to *PCCB* and *FADS1* expression, respectively, was then utilised to perform a phenome-wide scan spanning over 10,000 GWAS of the effect of expression of these two genes using SNP effect sizes from the IEUGWAS database (Supplementary Tables 11-13). Firstly, we found that increased cortical expression of *PCCB*, which was associated with deceased odds of SZ from a previous GWAS, was also linked to a reduction in other psychiatric phenotypes from self-reported UK Biobank GWAS such as worry, neuroticism, nervousness, and tenseness, supporting the utility of a *PCCB* agonist, like biotin, as a repurposing candidate. Secondly, the phenome-wide data for increased cortical *FADS1* expression demonstrated, as expected, a strong effect on lipids, including increased HDL and decreased triglycerides. While another candidate for BIP (*MAP2K2*) was suggested as a *trans*-pQTL by this approach, it was excluded to retain the most biologically confident associations. Although the MR approach did not add any additional candidate directional anchor genes (after exclusion of *MAP2K2*), it provided more support to *PCCB* and *FADS1*. A less conservative MR paradigm in terms of IV selection would likely yield more genes but as our TWAS/PWAS analyses were already discovery focused, we believe this would not be appropriate given the underlying assumptions of MR. We summarise the candidate directional anchor genes in table 1.

### Interaction networks related to directional anchor genes capture biology associated with schizophrenia and bipolar disorder

We sought to define a network of genes that display high confidence interactions with each candidate directional anchor gene using data from the STRING database, such that we can then construct a *pharmagenic enrichment score* using variants annotated to these genes. The number of direct interactions identified for each of the six candidate genes, excluding the gene itself, were as follows (Supplementary Tables 14): *CACNA1C* network (83 genes), *FADS1* network (16 genes), *FES* network (37 genes), *GRIN2A* network (54 genes), *PCCB* network (26 genes), and *RPS17* network (254 genes). All of these networks displayed significantly more interactions than what would be expected by chance alone (*P* < 1 × 10^−16^, Supplementary Table 15), with an example of two of these networks (*CACNA1C* and *FADS1*) visualised in Figure 3a. These networks likely represent heterogenous biological processes in which the directional anchor gene may participate, and thus, we sought to better understand the biology of these interacting genes by testing their overrepresentation within biological pathways and other ontological gene-sets. The six directional anchor gene networks each displayed overrepresentation in pathways related to the known biology of the candidate gene (Supplementary Tables 16-21). For instance, the *CACNA1C* network genes were enriched within several hundred gene-sets, many of which related to neuronal calcium channel biology along with systemic processes known to involve calcium signalling such as pancreatic insulin secretion. Furthermore, the *FADS1* network genes displayed an overrepresentation in several lipid and other metabolic related pathways, whilst *GRIN2A* network genes demonstrated a strong link to neuronal biology.

**Figure 3.**
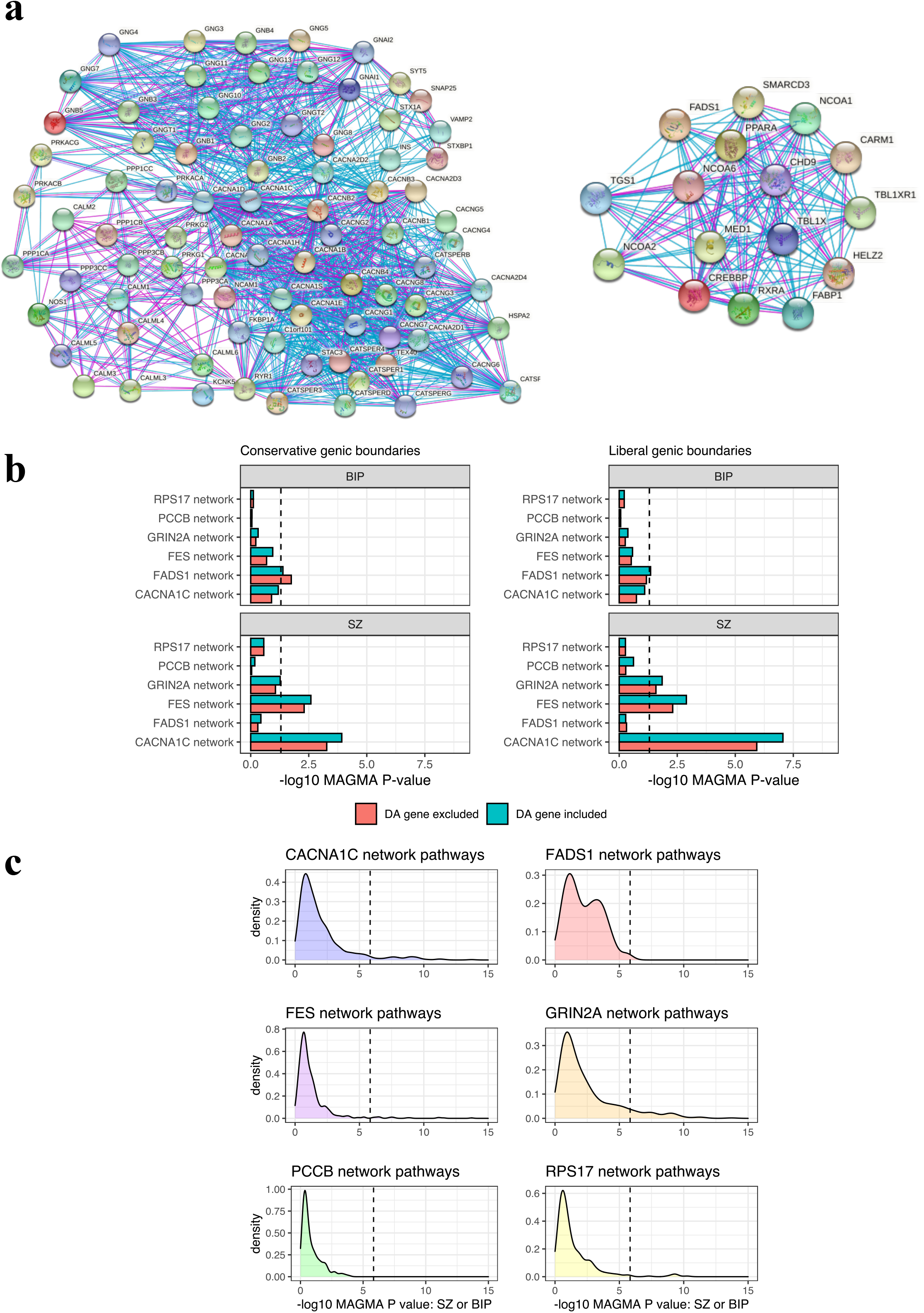
Biological networks interacting with candidate directional anchor genes. **(a)** Visualisation of two networks of genes that putatively interact with *CACNA1C* (left) and *FADS1* (right) based on experimental and curated database evidence. Blue edges represent evidence from curated databases, whilst purple edges denote experimentally determined evidence. (**b**) Gene-set association (MAGMA) of the entire network for each candidate directional anchor (DA) gene, with and without the DA gene included from the model. Dotted line represents nominal significance (*P* < 0.05). The MAGMA *P* value is derived from a model which tests whether the common variant signal within genes in the network is greater than what is observed amongst all remaining genes. Two genic boundaries were utilised to annotate SNPs to genes from the GWAS: conservative (5kb upstream, 1.5 kb downstream, left panel) and liberal (35 kb upstream, 10 kb downstream, right panel). (**c**) Kernel density estimation plots of the MAGMA gene-set association *P* value for each gene-set tested using either schizophrenia or bipolar results, whichever was more significant, which had a significant overrepresentation of genes within that network. The dotted line represents the Bonferroni significance level for approximately 34,000 gene-sets considered in the full analysis of all gene-sets that were tested for overrepresentation.

We then tested whether there was enrichment of the common variant signal for SZ or BIP in any of these networks, with and without the directional anchor gene included, using MAGMA (Figure 3b, Supplementary Table 22). The *CACNA1C* network was strongly associated with SZ (*P* = 8.87 × 10^−8^), even after removing *CACNA1C* itself (*P* = 1.19 × 10^−6^). The *FES* and *GRIN2A* networks demonstrated a nominal enrichment of the SZ common variant signal relative to all other genes, *P* = 1.28 × 10^−3^ and *P* = 0.014, respectively, remaining significant upon removing the relevant directional anchor genes. None of the other networks were associated with SZ when considering all genes, with only the *FADS1* network demonstrating a nominal association with BIP (*P* = 0.04). Given that these networks represent several different biological processes, we further hypothesised that specific gene-sets for which they were overrepresented may specifically display a stronger association with SZ or BIP (Supplementary Tables 23-28). Indeed, we show that all of the networks had at least one overrepresented pathway that was associated with SZ or BIP using Bonferroni (FWER < 0.05) and Benjamini-Hochberg (FDR < 0.05) correction, with the exception of the sets enriched in the *PCCB* network that only survived correction using FDR. Kernel density estimation plots of the MAGMA gene-set association *P* values are visualised in figure 3c, which show pathway-associations reaching these thresholds. We briefly describe the results for the *CACNA1C* and *GRIN2A* networks below for illustration. Pathways overrepresented in the *CACNA1C* network related to calcium channel activity displayed strong association with SZ, for instance, *voltage gated calcium channel process* (*P* = 2.80 × 10^−10^, *q* = 6.83 × 10^−7^), whilst the *regulation insulin secretion* pathway that also was enriched in the network was associated with SZ and trended towards surviving multiple testing correction for BIP. *GRIN2A* network members also displayed an enrichment amongst several neuronal pathways strongly associated with SZ, such as *synaptic signalling* (*P* = 3.88 × 10^−8^, *q* = 2.82 × 10^−5^). Taken together, these results suggest that pathways in which genes in each network participate are associated with psychiatric illness and reinforces the biological salience of these networks.

### Directional anchor gene network *pharmagenic enrichment scores* display significant trait associations after adjustment for genome-wide polygenic risk score

*Pharmagenic enrichment scores* (PES) were then constructed for the genes in each directional anchor gene network using SNP weights for SZ and BIP, respectively. SZ and BIP PES were considered for all six networks given the high genetic correlation between SZ and BIP, as well as extensive phenotypic overlap. We defined a training set of SZ (N = 631) and BIP cases (N = 1161) in the UK Biobank, with double the number of controls randomly, and independently, selected from individuals with no self-reported mental health conditions for each training set. Two methods were utilised to find the most parsimonious PES profile for each network, along with a genome-wide PRS for SZ and BIP - clumping and thresholding (C+T), and penalised regression (Table 2).

**Table 2.** Characteristics of the best performing schizophrenia and bipolar genome wide PRS along with a *pharmagenic enrichment score* for each directional anchor gene network.

| Score | N<sub>SNPs</sub><sup>1</sup> | Beta (SE)<sup>2</sup> | P | R<sup>2</sup> | Type<sup>3</sup> |
| --- | --- | --- | --- | --- | --- |
| *Bipolar disorder* |  |  |  |  |  |
| PRS | 1096096 | 0.62 (0.04) | 2.91x10<sup>-51</sup> | 4.96% | Penalised regression |
| *CACNA1C* network | 396 | 0.11 (0.04) | 2.97x10<sup>-3</sup> | 0.16% | Penalised regression |
| *FADS1* network | 196 | 0.06 (0.04) | 0.1 | 0.05% | C+T |
| *FES* network | 5 | 0.10 (0.04) | 4.87x10<sup>-3</sup> | 0.14% | Penalised regression |
| *GRIN2A* network | 143 | 0.17 (0.04) | 3.81x10<sup>-6</sup> | 0.39% | Penalised regression |
| *PCCB* network | 520 | 0.14 (0.04) | 2.22x10<sup>-4</sup> | 0.25% | Penalised regression |
| *RPS17* network | 13724 | 0.15 (0.04) | 1.07x10<sup>-4</sup> | 0.28% | Penalised regression |
| *Schizophrenia* |  |  |  |  |  |
| PRS | 1861450 | 1.01 (0.07) | 2.65x10<sup>-51</sup> | 9.55% | Penalised regression |
| *CACNA1C* network | 128 | 0.09 (0.05) | 0.1 | 0.09% | C+T |
| *FADS1* network | 36 | 0.04 (0.05) | 0.4 | 0.02% | C+T |
| *FES* network | 141 | 0.17 (0.05) | 1.41x10<sup>-3</sup> | 0.32% | C+T |
| *GRIN2A* network | 5037 | 0.18 (0.05) | 9.23x10<sup>-4</sup> | 0.35% | Penalised regression |
| *PCCB* network | 2393 | 0.07 (0.05) | 0.16 | 0.06% | Penalised regression |
| *RPS17* network | 77 | 0.15 (0.05) | 3.53x10<sup>-3</sup> | 0.27% | Penalised regression |

<sup>1</sup>SNPs with a non-zero coefficient after reweighting in the penalised regression model or independent SNPs after linkage disequilibrium clumping and thresholding (C+T).
<sup>2</sup>Bipolar disorder or schizophrenia log odds per standard deviation increase in the score (standard error)
<sup>3</sup>The two models evaluated were clumping and thresholding (C+T) or penalised regression (as implemented by the lassosum package)

#### Schizophrenia

In the SZ cohort, there were three network SZ PES which were significantly associated with increased odds of SZ after multiple testing correction including networks for *FES, GRIN2A*, and *RPS17* (Table 2, Supplementary Figure 1a). In the *GRIN2A* network PES featuring 5037 variants constructed using penalised regression, explained approximately 0.35% of phenotypic variance on the liability scale (OR per SD in score = 1.19 [95% CI: 1.09, 1.29], *P* = 9.23 × 10^−4^). We then conservatively adjusted for the best performing genome-wide SZ PRS and found that the *GRIN2A* network PES remained significantly associated with SZ. In the *FES* and *RPS17* networks, their PES were just below the threshold for significance after PRS adjustment (Supplementary Table 29). It is notable that the SZ network PES profiles were only marginally correlated with genome-wide SZ PRS (all *r* < 0.11, Supplementary Figure 2a), which suggests these scores may capture biologically aggregated risk which is distinct from the undifferentiated genome-wide signal. Interestingly, the majority of individuals with SZ (53.72%) had at least one elevated PES (for the disorder) in the highest decile, which was significant even after covariation for genome-wide PRS – OR = 1.45 [95% CI: 1.22, 1.67], *P* = 1.57 × 10^−3^. Interestingly, amongst individuals in this cohort with relatively low SZ PRS (lowest decile), 12 out of the 19 SZ cases had an elevated PES (63%), with a nominally significant association remaining between elevated PES and SZ amongst those with low genome wide PRS (*P* = 0.027). Upon considering only SZ cases in terms of low PRS, we found that 46.88% had at least one elevated PES. Taken together, these data suggest that some individuals with otherwise low SZ PRS have localised genetic risk within these biological networks. Given the relatively small number of SZ cases in the UKBB, we sought to replicate our results using an independent case-control cohort from the ASRB (N_Cases_ = 425, N_Controls_ = 251) rather than splitting the UKBB cohort into a training and validation set. The PES and PRS models were retrained in the UKBB from the same GWAS with the ASRB cohort removed. We were able to nominally replicate the association of the *FES* network PES with SZ in the ASRB (OR per SD = 1.21 [95% CI: 1.04, 1.38], *P* = 0.024), whilst the observed association between the *GRIN2A* and *RPS17* network PES and SZ and in the UKBB was not replicated (Supplementary Table 30).

#### Bipolar disorder

BIP PES within these networks was then profiled in the UKBB training set (Table 2, Supplementary Figure 1b). Interestingly, there were more of the directional anchor gene network PES associated with BIP than SZ, which may have been a reflection the greater statistical power afforded by the larger number of BIP cases in the UKBB. Specifically, all of the network BIP PES were significantly higher in cases, with the exception of the *FADS1* network PES for which there was only a trend towards significance. Analogous to the SZ cohort, the *GRIN2A* network PES explained the most phenotypic variance on the liability scale (0.39%), with each SD in the score associated with a 19% [95% CI: 12%, 26%] increase in the odds of BIP. Moreover, adjustment for BIP genome wide PRS did not ablate the significance of the *GRIN2A* network, *RPS17* network PES, and *FES* network PES, whilst the *PCCB* network PES trended towards significance (*P* = 0.1) after PRS covariation (Supplementary Table 31). The correlations between each PES and BIP PRS were also small (Supplementary Figure 2b), however, the *RPS17* network PES (*r* = 0.13), *CACNA1C* network PES (*r* = 0.14), and *PCCB* network PES (*r* = 0.13) were slightly larger in terms of their PRS correlation than what was observed for the SZ scores. We then investigated the characteristics of individuals with elevated BIP PES and found like SZ that almost half of the BIP cases (49.1%) had at least one PES greater than or equal to the 90^th^ percentile. There was also enrichment of BIP cases amongst participants with an elevated PES after adjusting for BIP PRS (OR = 1.19 [95% CI: 1.04, 1.34], *P* = 0.027). Considering BIP cases in the lowest decile of the BIP PRS distribution, 36% of them had at least one top decile PES despite their low genome-wide burden, although unlike SZ the association between elevated PES and case-status in this subcohort was not statistically significant. An independent BIP case-control cohort from the UKBB was utilised to attempt to replicate these associations (Supplementary Table 32), and we found that the network *RPS17* PES was significantly enriched in BIP cases in this validation cohort: whilst there was a trend for the *GRIN2A* network PES (*P* = 0.052).

#### Sensitivity analyses for the GRIN2A network PES

The *GRIN2A* network PES explained the most phenotypic variance for SZ and BIP, and survived covariation for genome-wide PRS – thus, we wanted to test whether constructing a PES for this network with the *GRIN2A* gene excluded would still be associated. In other words, we investigated whether there was an effect from variants mapped to the network without the directional anchor gene itself. For example, the *GRIN2A* network PES with the *GRIN2A* gene removed, was still significantly enriched in both SZ and BIP (*P*_SZ_ = 9.23 × 10^−4^ and *P*_BIP_ = 3.81 × 10^−6^). The relationship between genome-wide PRS and this PES was also examined in further detail by constructing a ‘residualised PES’ whereby we obtained the normalised residuals from a model that regressed SNP derived principal components, genotyping batch, and genome-wide PRS for BIP and SZ, respectively, on the *GRIN2A* network PES for either disorder. We posit that the individuals with an elevated residualised PES are more likely to represent true enrichment in that network given that the effect of the genome wide PRS, along with variables related to technical artefacts and population stratification, have been adjusted for. Encouragingly, we find that the correlation between the raw *GRIN2A* network PES for either disorder and their respective residualised PES were highly concordant, with the majority of individuals with an elevated *GRIN2A* PES (≥ 90^th^ percentile) also in that same quantile for the residualised PES (Figure 4a-b).

**Figure 4.**
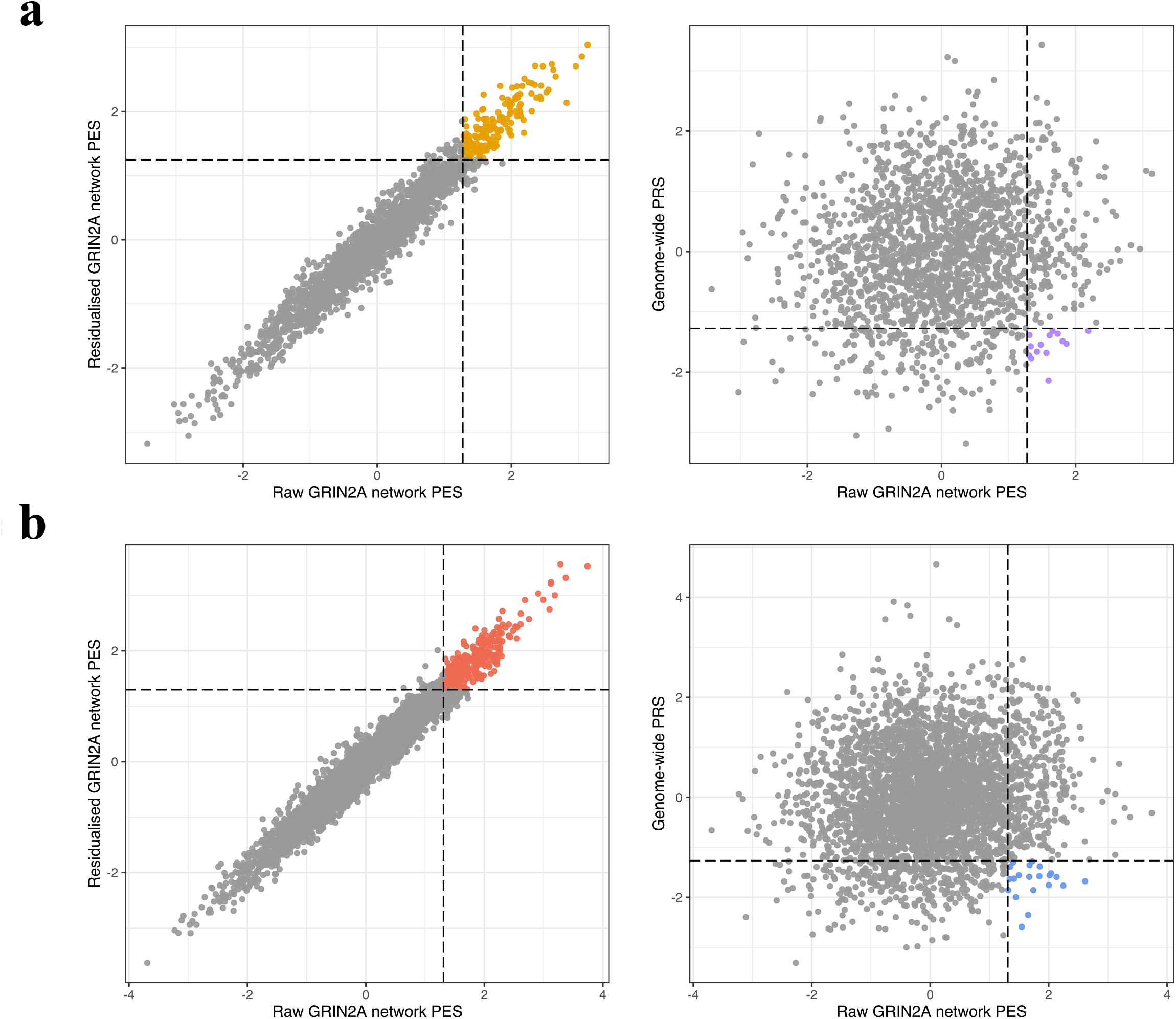
Schizophrenia and bipolar disorder *GRIN2A* directional anchor gene network *pharmagenic enrichment scores* and their relationship with PRS. The scatter plots denote the concordance between the scaled unadjusted (raw) *GRIN2A* network PES for SZ (**a**) and BIP (**b)** and both a residualised score and genome wide PRS. Specifically, the left-most scatterplots visualise the relationship between the raw *GRIN2A* network PES and the residuals from a model which regressed genotyping batch, ten SNP derived principal components, and genome wide PRS for the disorder in question (Residualised *GRIN2A* PES). The dotted lines represent the 90^th^ percentile of the raw PES and residualised PES, respectively. The points coloured orange (SZ) and red (BIP) indicate individuals with a PES in the 90^th^ percentile or above for both the raw and residualised scores. The right scatterplots plot the relationship between genome wide PRS for SZ or BIP and the *GRIN2A* network PES. In these instances, the dotted vertical line denotes the 90^th^ percentile of the *GRIN2A* PES, whilst the horizontal dotted line denotes the 10^th^ percentile of genome wide PRS. As a result, the points coloured purple and blue in the SZ and BIP plots, respectively, are individuals with low relative genome wide PRS (lowest decile) but high *GRIN2A* PES (highest decile).

### Phenotypic correlations of directional anchor gene network scores support their biological relevance

We then investigated the association of the directional anchor gene PES in an independent subset of the UKBB with other mental health phenotypes and systemic biochemical measures (Supplementary Tables 33-35 Figure 5). The correlation profile of each PES relative to these phenotypes may also support its clinical utility, whilst it also provides an opportunity to further establish what distinct properties these scores have from a genome wide PRS. Firstly, all SZ and BIP network PES, along with their respective PRS, were regressed against 33 blood and urine measures in up to 70,625 individuals, whilst oestradiol in females was also additionally considered in a sex stratified analysis (Figure 5a). In both sexes, we found that the *FADS1* network PES for SZ and BIP was significantly correlated with lipid related traits after conservative Bonferroni correction for all PES/PES-trait pairs tested (*P* < 1.11 × 10^−4^). For instance, these *FADS1* network PES were negatively correlated with HDL cholesterol and apolipoprotein A1 levels, whereas increase in the same PES was associated with higher measured triglycerides. The *FADS1* network PES were also significantly associated other non-lipid biochemical traits including alkaline phosphatase, sex-hormone binding globulin (SHBG), and urate. Notably, after adjusting PES for a genome-wide PRS for SZ or BIP, did not ablate its association with the disorder, suggesting that these signals are not a product of genome-wide polygenic inflation (Supplementary Tables 36-39). To the contrary, there was evidence that PRS was correlated in the opposite direction with lipids to that of the corresponding PES. Given the strong lipid related signals, we also adjusted for statin use in an additional sensitivity analysis, but this similarly did not markedly impact the findings (Supplementary Table 39). Using less stringent FDR correction (FDR < 0.05), revealed more PES association with biochemical measures. This included a negative correlation between both the *PCCB* and *FES* network PES for SZ and insulin-like growth factor 1 (IGF1); as well as positive correlation between *RPS17* network PES for SZ and creatinine. However, there was no direct effect of SZ or BIP PRS on IGF-1 or creatinine, with FES-related tyrosine kinase activity previously shown in the literature to be associated with IGF-1 biology [47]. Sex stratified analyses identified even more PES associated with a biochemical trait (Supplementary Table 34) – for example, in males the SZ *PCCB* network PES was positively correlated with SHBG, which interestingly is in the opposite direction to the correlation of SHBG observed with the *FADS1* network PES, further highlighting biological heterogeneity amongst different networks. The BIP *CACNA1C* network PES in males was also positively correlated with direct bilirubin using an FDR cut-off, whilst the BIP *GRIN2A* network PES was negatively correlated with measured total protein. Finally, we formally tested for evidence of sexual dimorphic effects of PES/PRS on each biochemical measure and revealed nominal evidence of heterogeneity between sexes in these associations for some traits such as the effect of the *CACNA1C* network PES on direct bilirubin (Supplementary Table 40).

**Figure 5.**
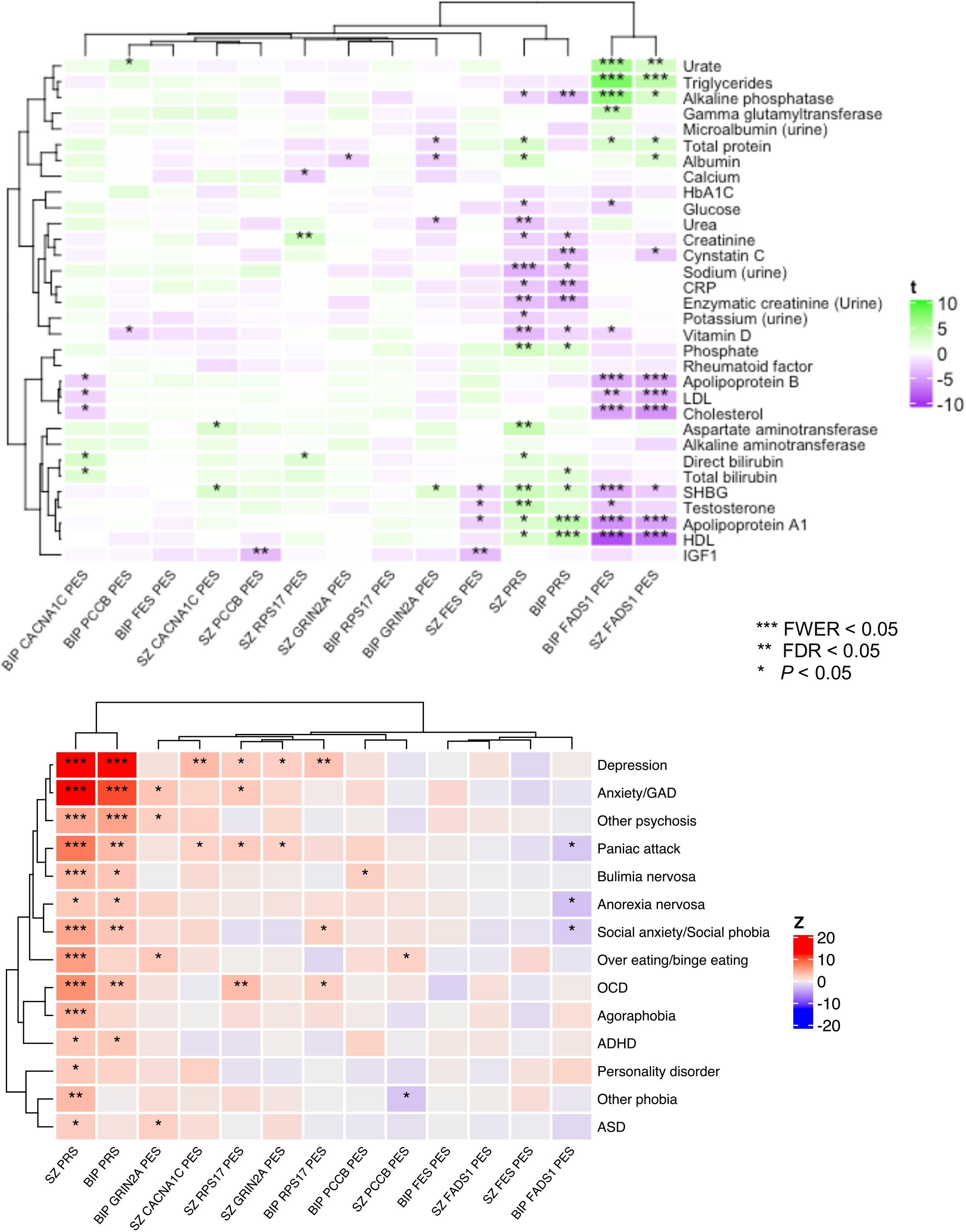
Phenome-wide association studies (pheWAS) of each network PES or PRS related to serum or urine biochemical measures and mental health disorders. Heatmap of the association between each network PES and PRS with each trait tested for the biochemical measures (top) and self-reported mental health disorders (bottom). Traits ordering derived from clustering by Pearson’s distance. The variable visualised in the heatmaps for the continuous biochemical traits was the regression *t* value (beta/SE), whilst for the binary mental health phenotypes it was the corresponding *Z* value from the logistic regression, whereby *Z* > 0 equates to an odds ratio for the disorder > 0. Asterisks were utilised to denote the significance of the association -* = *P* < 0.05, ** = false discovery rate (FDR) > 0.05, and *** = family-wise error rate (FWER) < 0.05.

We also performed a phenome-wide association study of each score with 14 self-reported mental health disorders in the UKBB cohort, excluding SZ and BIP (Figure 5, Supplementary Table 35). The number of cases ranged from 66 for attention deficit/hyperactivity disorder (ADHD) to 22,974 for depression, with the same cohort of 70,625 individuals without a self-reported mental health condition not featured in the SZ or BIP training/validation sets leveraged as controls. Unsurprisingly, we found that SZ and BIP PRS were strongly associated with increased odds of several mental health disorders after Bonferroni correction, but we also found network PES associated with some of these phenotypes using FDR < 0.05 as the multiple-testing correction threshold. Specifically, there was an association between the SZ *CACNA1C* PES and increased odds of depression, whilst the SZ *RPS17* network PES was associated with increased odds of self-reported OCD. These disorders were also associated with elevated SZ PRS, however, both PES remained significantly higher in those with the respective self-reported phenotypes even after covariation for the effect of the SZ PRS. There were also several other nominal associations (*P* < 0.05), including one of particular interest in the case of the BIP *FADS1* network PES, for which a higher score displayed some evidence of a protective effect on self-reported anorexia nervosa. Whilst this association does not survive multiple testing correction, and thus should be interpreted cautiously, it is notable as the *FADS1* network PES was associated with lipid profiles in an analogous direction to what has previously shown to be genetically correlated with anorexia nervosa GWAS via LD score regression [11, 21]. In summary, these data coupled with the biochemical associations support the distinct nature of network PES from PRS and emphasise the unique insights that can be afforded by these partitioned scores.

## DISCUSSION

In this study, we leverage genetics to identify novel drug repurposing candidates for psychotic disorders and show how they may be directed specifically to patients. This precision medicine approach is critical given that phenotypic and genetic heterogeneity confounds traditional interventions designed to target the entire disease population. We believe that a key advance in this study is that it provides a direct link between compounds with evidence for efficacy at the population level, a putative expression related mechanism, and genetic risk scores in the network of genes that interact with the prioritised drug target.

Transcriptomic and proteomic data integrated with GWAS through GReX (TWAS/PWAS) and causal inference (MR) revealed six interesting target genes for SZ or BIP that could be modulated in a risk decreasing direction by an approved drug. Compounds which target *GRIN2A* in the risk decreasing direction, one of the genes prioritised for SZ, have previously been subjected to randomised control trials as an adjuvant to antipsychotic treatment. Specifically, *N*-acetylcysteine and glycine intervention studies suggested that these compounds could be effective in improving multiple symptom domains including negative and cognitive related dimensions of the disorder [50, 51]. Omega-3 fatty acids, which are related to the biology of the BIP candidate gene *FADS1*, have also been of interest in that disorder, although trials have had contradictory results in terms of benefit [52]. This heterogeneity is unsurprising given the complexity of the BIP phenotype and demonstrates the utility of an approach such as ours which could more specifically target these interventions. The remaining genes and respective repurposing candidates all had plausible biological links to neuronal biology or psychiatric illness [53–56], and thus, warrant further investigation of their utility even without genetic stratification.

While these six candidates were supported by genetically regulated mRNA expression this was not confirmed by genetically regulated protein expression, however, this was probably due to reduced power to detect proteome related associations as this data is still relatively limited [48, 49]is still notably. There are also some limitations to using both the GReX and MR frameworks for target identification [7, 43, 46], although these are mitigated by a suite of sensitivity analyses we performed, including probabilistic finemapping and colocalization, which strengthen our confidence in these six genes. Ideally, future statistical and molecular study of these association signals should be undertaken to refine our understanding of the role of these genes in the pathophysiology of SZ and BIP.

We outlined a mechanism whereby genetic risk for the disorder could be profiled amongst the network of genes which were prioritised as repurposing opportunities and act as candidate directional anchor genes based on their direction of effect (network PES). Crucially, we observed a notable portion of cases with low overall PRS but at least one elevated PES, further supporting the biologically unique insights that may be gained from PES relative to an undifferentiated genome wide approach. Most of the scores also considered demonstrated at least nominal significance for association with either SZ or BIP, with PES like the *GRIN2A* network PES significantly enriched in both SZ and BIP even after conservatively correcting for the effect of a genome wide PRS. However, it should be noted that the training sets we used in this study were modestly powered, and larger training sets would be beneficial given that partitioned scores like PES will have smaller overall effect sizes than PRS. The relatively small trait-effect sizes of PES in terms of their phenotypic variance explained does not also necessarily preclude individual level relevance, particularly because PES like the *FADS1* network PES displayed strong correlations with traits relevant to that network such as measured lipids. Penalised regression was also applied to the construction of these network PES rather than just clumping and thresholding as was undertaken in previous PES studies [18–20]. These penalised regression PES did explain more variance in most of the networks considering the training set compared to clumping and thresholding, although selecting the appropriate constraint and tuning parameters when many combinations perform similarly is an ongoing challenge for such approaches. For example, the best performing *FES* network PES for BIP was derived through penalised regression, however, it only included five variants, which is not representative of the overall network. These issues highlight the need for future study in the construction of PES, particularly as popular genome wide approaches for PRS like LDpred and SBayesR would need to be methodologically adapted for a local gene-set or network implemention [15]. The proportion of phenotypic variance explained by PES derived using these networks could also be boosted in future by incorporating rare and structural variation, as well as re-weighting effect sizes informed by functional annotation.

While there is still work needed to confirm that the PES can effectively triage an individual’s suitability for drug repurposing candidates in psychiatry and other disorders, this study represents a key methodological advance as it predicts the desired direction of effect needed to modulate a given target within the context of a PES. Previously, it was not clear whether agonism or antagonism of genes in the set would be clinically useful. Randomised placebo control trials of the PES approach would be a further step to demonstrate its utility and could involve repurposed drugs stratified by the relevant PES or more complex study designs, such as the multi-crossover “*N*-of-one” approach [57, 58]. In summary, we present a novel framework to inform precision drug repurposing in psychiatry that could account for individual level heterogeneity in genetic risk factors, and therefore, improve patient outcomes.

## Data Availability

The analysis code for this study can be found in the following GitHub repository: https://github.com/Williamreay/Directional_anchor_gene_psyciatric_PES. Researchers can access the full UK Biobank data upon approval (https://www.ukbiobank.ac.uk/enable-your-research/apply-for-access). The ASRB cohort is available upon reasonable request and ethics approval (https://www.neura.edu.au/discovery-portal/asrb/). The remaining data are all publicly available, as outlined in the studies cited at relevant positions throughout the main text.

https://www.neura.edu.au/discovery-portal/asrb/

## ACKNOWLEDGEMENTS

This work was supported by a National Health and Medical Research Council (NHMRC) grant (1188493). M.J.C. is supported by an NHMRC Senior Research Fellowship (1121474), URL: https://www.nhmrc.gov.au/. The funders had no role in study design, data collection and analysis, decision to publish, or preparation of the paper. W.R.R was supported by an Australian Government Research Training Program Stipend. This research was conducted using the UK Biobank resource under the application 58432.

## AUTHOR CONTRIBUTIONS

W.R.R. conceived and designed the study with input from M.J.C. W.R.R. performed the primary analyses. M.P.G assisted with the quality control and processing of the UK Biobank cohort. J.R.A performed the SNP imputation in the ASRB cohort. M.J.C, V.J.C, and M.J.G contributed the ASRB cohort, as well as to critical interpretation of the results and drafting of the manuscript.

