## Supplementary Materials for "Directional anchor genes refine polygenic informed treatment selection in schizophrenia and bipolar disorder"

**SUPPLEMENTARY TEXT AND FIGURES**

**FUSION implementation of transcriptome and proteome wide association studies (TWAS/PWAS)**

*TWAS*

Brain related SNP weights (multivariate GReX) TWAS were derived from GTEx v7 and PsychENCODE, whilst whole blood weights were also obtained from GTEx v7 [1, 2]. The GTEx v7 SNP weights comprise data from 11 different brain regions – with the sample sizes of the cohorts utilised for GReX estimation as follows: amygdala (N=88), anterior cingulate cortex (N=109), caudate (N=144), cerebellar hemisphere (N=125), cerebellum (N=154), cortex (N=136), frontal cortex (N=136), hippocampus (N=111), hypothalamus (N=108), nucleus accumbens (N=130), and putamen (N=80). We implemented FUSION using default parameters for these 11 tissues using the HapMap3 SNPs from the 1000 genomes phase 3 European reference panel as a linkage disequilibrium (LD) estimate, to correspond with how the weights were calculated with those same HapMap3 SNPs. The TWAS using whole blood weights utilised the same LD reference strategy, with the number of GTEx v7 participants in these models 369. Frontal or cerebral cortex tissue from the larger PsychENCODE cohort was also utilised in terms of SNP weights, with a sample size of 1695. As described in the original PsychENCODE publication, gene-wise GReX were estimated only all imputed SNPs, not just the HapMap3 panel, and thus, we utilised the full suite of the phase 3 1000 genomes European subset as the LD reference. Summary statistics were also munged using the FOCUS wrapper for the ‘munge_sumstats.py’, whereby SNPs were retained with an imputation INFO > 0.9, as well as removing indels, strand ambiguous SNPs and SNPs with MAF < 0.01. In total, considering all GReX models that survived the FUSION pipeline’s internal QC before calculating the TWAS *Z*, there were 55,834 and 54,986 models tested for SZ and BIP, respectively. This equated to approximately 18,000 unique genes tested for both disorders. However, we conservatively adjusted for all of the models tested in terms of multiple-testing correction to only priortise the most statistically significant signals for further consideration.

*PWAS*

There were two tissues for which were obtained SNP weights related to protein expression – the dorsolateral prefrontal cortex (DLPFC, N = 376) and plasma (N = 7213) [3, 4]. It should be noted that the plasma weights were derived from the European subset of the study cohort, however, the authors only used the elastic net method to derive GReX. Analogous to the difference between the GTEx v7 and PsychENCODE studies above, the DLPFC SNP weights were estimated using the HapMap3 panel, and thus, we only used those SNPs as an LD reference. The plasma PWAS utilised the full reference panel. The number of protein models available in each tissue for the two GWAS was as follows: SZ – plasma = 484, DLPFC = 1443. BIP – plasma = 350, DLPFC = 1400.

**Probabilistic finemapping of TWAS signals which represent a plausible repurposing opportunity (FOCUS)**

We sought to finemap the region of genes for which their genetically regulated expression was associated with SZ or BIP after Bonferroni correction in a fashion that could be counteracted by an approved drug [5]. There were two reference panels utilised for finemapping – for genes uncovered from a GTEx v7 tissue, we utilised the default combined FOCUS SNP weight set which collated GTEx v7 tissues, DLPFC (CommonMind), blood (YFS, NTR), and adipose (METSIM) SNP weight sets (<https://www.dropbox.com/s/ep3dzlqnp7p8e5j/focus.db?dl=0>), with genes discovered using the PsychENCODE weights finemapped specifically using that panel given the different LD parameters and its more complete set of genes with cis-heritable models in that one tissue. The default FOCUS panel was tested by prioritising brain related GReX, followed by predictive accuracy in the absence of a brain tissue SNP-weight set. In SZ, there were twelve such candidates with four genes displaying a high posterior inclusion probability (*PIP*) of over 80% of being in the 90% credible set (*GRIN2A*, *FES, PCCB,* and *CACNA1D*), whilst two additional genes surpassed the more relaxed criterion of *PIP* > 0.4 (*CACNA1C* and *RPS17*). The remaining genes all had very low *PIP*, which suggested they were not a likely causal gene at that locus based on TWAS data (*NEK4*, *MAPK3*, *TLR9*, *CHRNA2, PSMB8, SERPINC1*, and *CTSS*). There were two candidate genes for BIP (*NEK4* and *FADS1*), with only *FADS1* surpassing the relaxed *PIP* threshold of 40% using the PsychENCODE SNP weights.

In the case of candidate gene achieving PIP > 0.8, we also considered the use of a more conservative Bernoulli prior for the models of each causal indicator [$c \sim\text{ Bern}(p)$] of *p* = 1 x 10^-5^, rather than the default *p* = 1 x 10^-3^, with the implemention of these priors outlined more extensively in Mancuso *et al.* [5]. We found this had no large effect on the *PIP* for three of the genes, with the updated *PIP* as follows: *FES* – *PIP* = 1, *GRIN2A*– *PIP* = 0.926, and *PCCB* – *PIP* = 1. The *CACNA1D* *PIP* was ablated upon using this more conservative prior (*PIP* = 0.478), with more evidence in this construct for *GNL3* as a candidate causal gene for this region (*PIP* = 0.88). As described in the following section, we believe that the complexity of the region in which *CACNA1D* is located and the weakness of its GReX model necessitates removing this gene as a candidate DA-gene.

*CACNA1D region*

Despite *CACNA1D* exhibiting a high *PIP* using the default prior, we believe that this association is not confident enough to carry forward with as a candidate directional anchor gene. Firstly, whilst this gene displayed a strong association in the PsychENCODE cortical TWAS (*Z* = 6.38), its GReX models only explained a very small amount of phenotypic variance that was only nominally statistically significant relative to the other GReX models considered as candidate DA-genes (cross-validated *R*^2^ = 0.003, *P* = 0.024). This GReX model, which performed best using a Bayesian sparse linear mixed model in the PsychENCODE study, was not found in any of the smaller sample size GTEx tissues – suggesting that the effect of *cis*-acting SNPs on expression was very weak for this gene and could only be detected at larger sample sizes. For comparison, the mean cross-validated variance explained in mRNA expression by GReX models of non-MHC genes from the PsychENCODE weight set was approximately 8.64%. We subjected the SNP weights in the *CACNA1D* GReX model and the same signal in the SZ GWAS to colocalisation analyses and found no strong evidence that *CACNA1D* expression was associated with both traits, either driven by a different or the same underlying causal variant (*PP*_H3_ = 0.164, *PP*_H4_ = 0.228), which is likely driven by the fact that the GReX model is only weakly statistically significant in terms of its correlation with expression. Finally, the region in which *CACNA1D* is located on chromosome 3 is a gene-dense region with multiple different biologically plausible causal genes such as *NEK4*, and *GNL3*, which becomes the most likely causal candidate using the more conservative prior, as outlined above.

**Selection of eQTL and pQTL as instrumental variables**

The selection of QTLs as instrumental variables (IV) can be challenging, particularly due to the onerous assumptions of MR for a valid IV. In most cases, only a single or a handful of IVs can be specified for mRNA or protein expression of any given gene, and, as a result, the suite of sensitivity analyses applied to more polygenic MR cannot be applied such as median, modal, and Egger regression based estimators [6, 7]. This reinforces that only the strongest QTL related signals should be chosen, as well as those associated with further genes such that there is less possibility for horizontal pleiotropy through the action of another transcript or protein. MR aims to infer a causal relationship and a corresponding causal estimate, which justifies conservatism in its approach. In the following section, we detail how IVs were chosen from the eQTL and pQTL studies we selected for analyses.

*Expression quantitative trait loci (eQTL)*

An eQTL study from blood (eQTLgen) and brain (MetaBrain) were selected as the source of eQTLs [8, 9]. These studies had larger sample sizes than the cohorts utilised to derive GReX models for TWAS, however, TWAS weights were unavailable in both instances. The eQTLgen study was a meta-analysis of blood-based eQTL cohorts, with a sample size up to 31,684, whilst MetaBrain was also a meta-analysis of brain eQTL cohorts across different regions. We selected three MetaBrain tissues which had a European ancestry sample size > 200, cortex (N = 2970), cerebellum (N= 492), and basal ganglia (N = 208). In both tissue types, blood, and brain, we sequentially applied the following quality control steps

i) Retain only variants associated with mRNA expression using a genome-wide significance threshold (*P* < 5 x 10^-8^). This threshold would be over-conservative for eQTL discovery, however, given IVs must be strongly associated with the exposure, we concluded that using only genome-wide significant signals would assist to define the strongest signals.
ii) Variants associated with three or fewer genes at genome-wide significance were retained to attempt to minimise the inclusion of highly pleiotropic signals or LD dense regions where many genes can be implicated.
iii) Identify the most significant independent signals (LD *r*^2^ < 0.001) from each 10000 kilobase clump (one megabase), with LD inferred using the 1000 genomes phase 3 reference panel. This is also conservative and reduces the number of genes with suitable IVs available – however, this assists to further refine the strongest signals in terms of statistical significance. It should be noted that statistical significance does not necessarily define the most likely causal variant, with future work required to refine likely causal candidates in eQTL studies with greater precision. As outlined in the main text, our TWAS was discovery orientated, meaning that these MR analyses could afford their conservatism.

The above quality control steps left the following numbers of genes to be tested in each tissue with a suitable IV – blood (N_Genes_ = 2880), cortex (N_Genes_ = 1833), cerebellum (N_Genes_ = 1162), and basal ganglia (N_Genes_ = 375).

*Protein quantitative trait loci (pQTL)*

As in the eQTL analyses we used both blood and brain pQTLs [3, 7]. The blood pQTL were assembled from the “tier 1” pQTLs from Zheng *et al.*, as outlined fully in that study. Briefly, pQTL results from five studies were collated and designated a level of confidence using a tiered system – whereby tier 1, the highest level of confidence, consisted of genome-wide pQTLs that were associated with less than five genes as well as exhibiting consistent effects (based on heterogeneity and colocalisation) across contributing studies they were available in. We further filtered these variants by retaining only those associated with three or fewer genes at genome-wide significance, as well as gene-wise filtering of any dependent SNPs (LD *r*^2^ > 0.001). The LD selection step for the pQTLs was slightly less stringent than what was applied to the eQTLs as it was performed per gene rather than per one megabase clump. This was because there were significantly fewer pQTL to begin with, and many genes did not have their protein levels assayed in the contributing studies. There were 891 genes remaining after the above analysis with a valid pQTL. A small number of *trans* pQTL were included in the tier 1 category, which we retained for discovery purposes but did not include any of these genes as final candidate directional anchor genes. The ROSMAP DLPFC cohort contributed the brain pQTL (N = 376), which was the same cohort utilised for estimating the PWAS weights. We also retained only genome-wide significant pQTLs that were also at least nominally significant (*P* < 0.05) in a smaller sample size replication cohort from the Banner Sun Health Research group (N = 189). Thereafter, SNPs were retained if they were associated with three or fewer genes, with the same gene-wise clumping applied as was the case in the blood pQTL studies. In total, there were 479 DLPFC genes with a valid pQTL.

**Sensitivity analyses for candidate target genes from Mendelian randomisation**

There were three candidate directional anchor genes from the MR analyses that could represent repurposing opportunities for schizophrenia (*PCCB*, *NEK1*, and *PTK2B*), whilst there were two for bipolar disorder (*FADS1* and *MAP2K2*). All of these genes were found using eQTLs as IVs, except for *MAP2K2*. Given that the *MAP2K2* IV was a *trans*-pQTL, we decided not to include this gene as a candidate directional anchor gene for a PES network, however, future study to refine the mechanism by which the IV SNP acts in *trans* to regulate protein expression could render this gene as a suitable repurposing opportunity. For the remaining genes we considered the following three factors as sensitivity analyses:

i) The genomic location of the IV SNP relative to the gene of interest. IVs located within or proximal to the gene of interest could be viewed as stronger evidence, although genic proximity does not guarantee causality.
ii) Colocalisation between expression of the gene and schizophrenia or bipolar disorder
iii) Phenome-wide MR (MR-pheWAS) using SNP effects from the IEUGWAS db. Specifically, we harmonised the IV effect allele to that of the other GWAS in the database and calculated a Wald ratio, along with its corresponding standard error and *P* value, to define other phenotypes for which expression of that gene could be causally implicated. These data were also helpful to refine other genes which could be associated with the IV.

It should be noted that all of the candidate genes only had a single IV, and, as a result, conventional polygenic MR sensitivity analyses like models with other IV assumptions (majority valid, plurality valid, and InSIDE) could not be implemented. We outline the results for the sensitivity analyses and MR-pheWAS for each gene below.

*PCCB*

The IV SNP for *PCCB* was an eQTL from the MetaBrain cortex meta-analysis (rs480330), and is intronic within the *PCCB* gene itself, supporting its relevance for that gene. Colocalisation analyses using default priors implicated that *PCCB* mRNA expression and schizophrenia were both associated, however, there was evidence of a different underlying causal variant (*PP*_H3_ = 0.95. Interestingly, if the prior probability for a shared causal variant is increased from the default of 1 x 10^-5^ to 1 x 10^-5^, there is a corresponding rise in the posterior probability of a shared underlying causal variant which approaches 40%. The MR-pheWAS revealed several interesting potential associations which support that increased cortical expression *PCCB* could be protective for schizophrenia, for instance, an association with several other psychiatric phenotypes as outlined in the main text. Expression of *PCCB* was linked to some circulating biochemical factors which could plausibly mediate some of its relationship with schizophrenia, such as sex-hormone binding globulin, testosterone, and bilirubin. Full results of these analyses are presented in supplementary table 11. Importantly, the strongest association was with *PCCB* expression, rather than other genes, with the *PCCB* eQTL data in the IEUGWAS database from blood.

*NEK1*

There was moderate evidence of a different underlying causal variant (*PP*_H3_ = 0.76, *PP*_H4_ = 0.24), although the evidence for H_4_ (shared causal variant) does trend towards 0.8 if you have a larger prior probability for that hypothesis > 1 x 10^-5^. The IV for this gene was intergenic and associated with three other genes at phenome-wide significance, including more significantly with the close-by gene *SH3FR1*, therefore, this association should be treated cautiously*.* Moreover, this gene did not survive correction in the TWAS but trended towards it *Z* = 3.43, *P* = 6.14 x 10^-4^. We decided that this signal could not be fully resolved given that the IV was associated with the expression of another proximal gene with more significance than *NEK1*.

*PTK2B*

There was strong evidence of a different underlying causal variant (*PP*_H3_ = 1), remaining consistent with a larger prior probability for H_4_. The IV SNP for this gene is intronic within the nearby *TRIM35* and the association of this gene with SZ via MR is larger, therefore, this is a lower confidence association with *PTK2B*.

*FADS1*

There was strong evidence of a different underlying causal variant between *FADS1* expression and bipolar, although both were associated (*PP*_H3_ = 1). The IV SNP is a non-coding transcript exon variant for the *FADS1* proximal *MYRF* gene, however, the MR-pheWAS supports a strong effect on lipid biology consistent with this gene and nearby *FADS2* (Supplementary Table 12)*,* as does the TWAS finemapping relative to *MYRF*. Given this gene also had support from TWAS, and a more biologically consistent MR-pheWAS result, we selected this as a candidate directional anchor gene.

**UK Biobank cohort sample specification and phenotype definition**

The UK Biobank is a large prospective cohort of approximately 500,000 participants for which extensive clinical, phenotypic, genetic, and molecular data are available [10]. Our group has previously processed the SNP array data through a series of quality control steps to retain a final cohort with 336,896 unrelated participants with white British ancestry for which 13,568,914 high quality (INFO > 0.8 or physically genotyped, missingness < 2%, no strong Hardy-Weinberg deviations) variants were available with a minimum minor allele frequency of 0.001%. Full details of the quality control applied are outlined in Reay *et al.* [11].

We utilised an integrative strategy for defining cases of schizophrenia and bipolar disorder. Given the diagnostic uncertainty and heterogeneity associated with these two disorders, we decided to implement a relatively broad phenotyping approach, however, given the discovery orientated nature of these analyses we feel that this is warranted. Specifically, we utilised three lines of evidence to define a schizophrenia or bipolar disorder diagnosis – self-reported doctor diagnosis at the assessment interview, linked primary or secondary ICD-10 inpatient records, and responses to the mental health questionnaire (MHQ) question regarding what mental illness a person had ever been diagnosed with by a psychiatrist. We selected individuals who satisfied any of these criteria and detail the specific phenotype codes below, note that individuals will satisfy multiple criteria in many instances, so the total is not a sum of each category, rather anyone who appears in at least one of the categories.

*Schizophrenia*:

- Self-reported at baseline (field 20002, code = 1289) – N = 334
- ICD-10 primary of secondary (field 41202 and 41205, codes = F20.1-9) – N = 453
- Mental health questionnaire (of 110,278 individuals in our genotype cohort who completed) – N = 90
- Total number of cases = 631

*Bipolar disorder:*

- Self-reported at baseline (field 20002, code = 1291) – N = 921
- ICD-10 primary of secondary (field 41202 and 41205, codes = F31.1-9) – N = 915
- Mental health questionnaire (of 110,278 individuals in our genotype cohort who completed) – N = 546
- N = 485 individuals self-reported at baseline and were hospitalized with a relevant ICD-10 code.
- Total number of cases = 1657

Controls were defined using the mental health questionnaire data also. These participants were 75,201 European, unrelated individuals with genotype data available which completed the MHQ and did not self-report any mental health conditions and did not meet any of the baseline SZ or BIP conditions. Controls were randomly selected from this cohort of 75,201 participants such that there were double the number to that of cases in the schizophrenia, bipolar disorder training, and bipolar disorder test set. The controls used in each of these three cohorts were independent of one another. In the case of the penalised regression (lassosum) models we used the controls from the opposite disorder as the LD reference, that is, for the schizophrenia cohort, the independent bipolar training cohort controls were used to account for LD in the regularization process. In the schizophrenia training cohort (N_Cases_ = 631, N_Controls_ = 1262) the schizophrenia cases were 63.2% male with a mean age of 55.4 (SD = 8.14), whilst the controls were majority female (51.8%) and older with a mean age of 56.6 (SD = 7.58). The BIP training set, which was 70% of the UKBB cases and double the number of randomly selected controls (N_Cases_ = 1161, N_Controls_ = 2322), had cases (56.8%) and controls (52%) which were majority female, a younger mean age of cases (55.6 vs 56.3). The composition of the BIP training set (N_Cases_ = 496, N_Controls_ = 992) was similar: cases – 56.3% female, mean age = 55.6 (SD = 8.11); controls – 51.6% female, mean age = 56.7 (SD = 7.63). In terms of genotyping batch, no more than 2% of any of the cohorts were from any one batch, however, batch was still included as a downstream covariate for stringency.

**Australian Schizophrenia Research Bank (ASRB) cohort**

The ASRB cohort in terms of genotyping and quality control has been described extensively previously [12]. Briefly, the ASRB was deeply phenotype cohort of recruited cases and non-neuropsychiatric controls as outlined by Loughland *et al.* [13]. The ASRB was used at the validation cohort for schizophrenia due to the comparatively small number of cases in the UKBB, however, given the ASRB was part of the schizophrenia GWAS, we retrained the models for replication in the UKBB training cohort with summary statistics that had the ASRB removed. After quality control, there were 425 cases (66.6% male, mean age = 39.88 (SD = 10.92) and 251 controls (56.2% female, mean age = 39.50 (SD = 13.4). There were 7,154,884 variants in this cohort available for analysis after post-imputation filtering.

**SUPPLEMENTARY FIGURES**

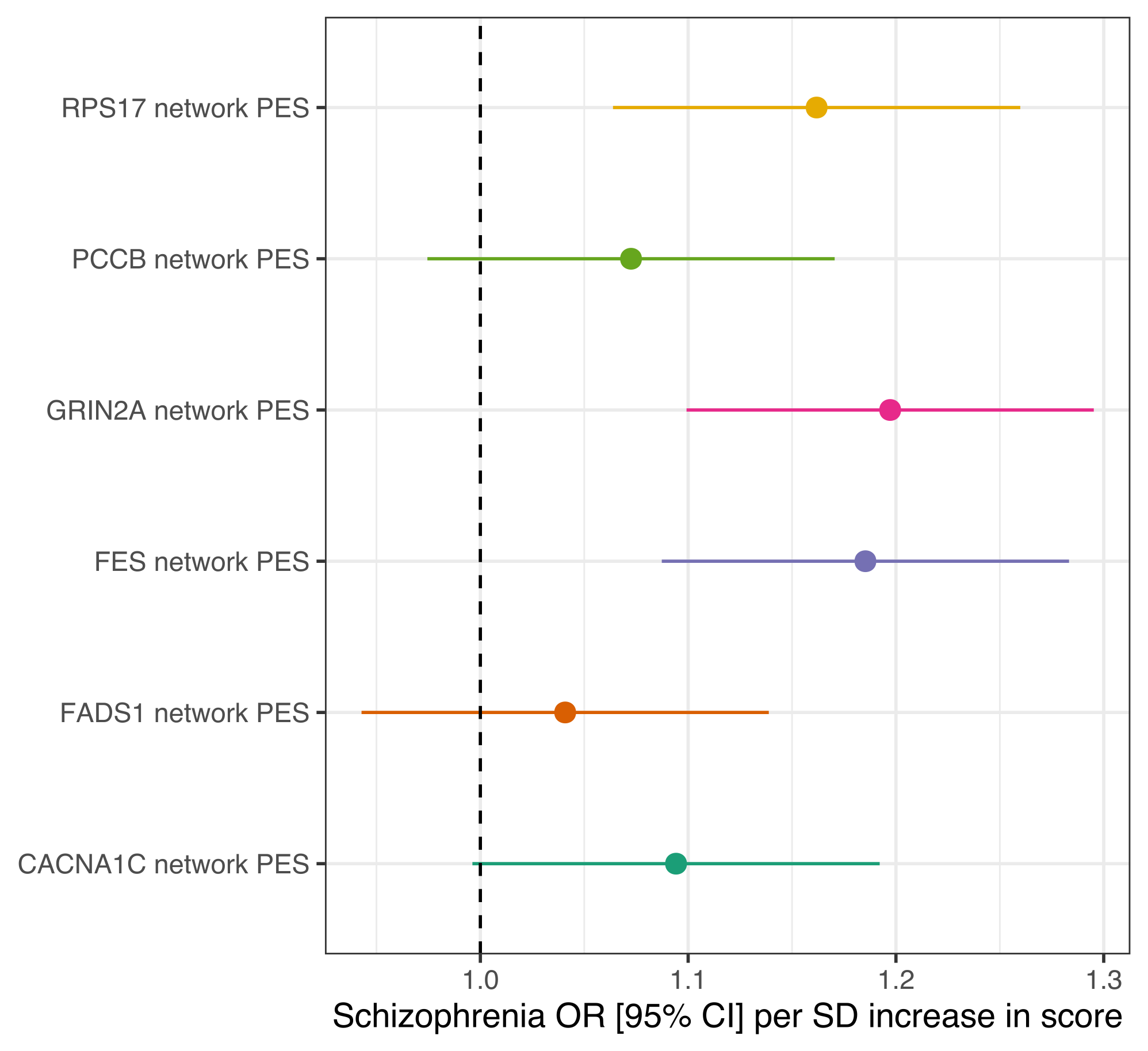

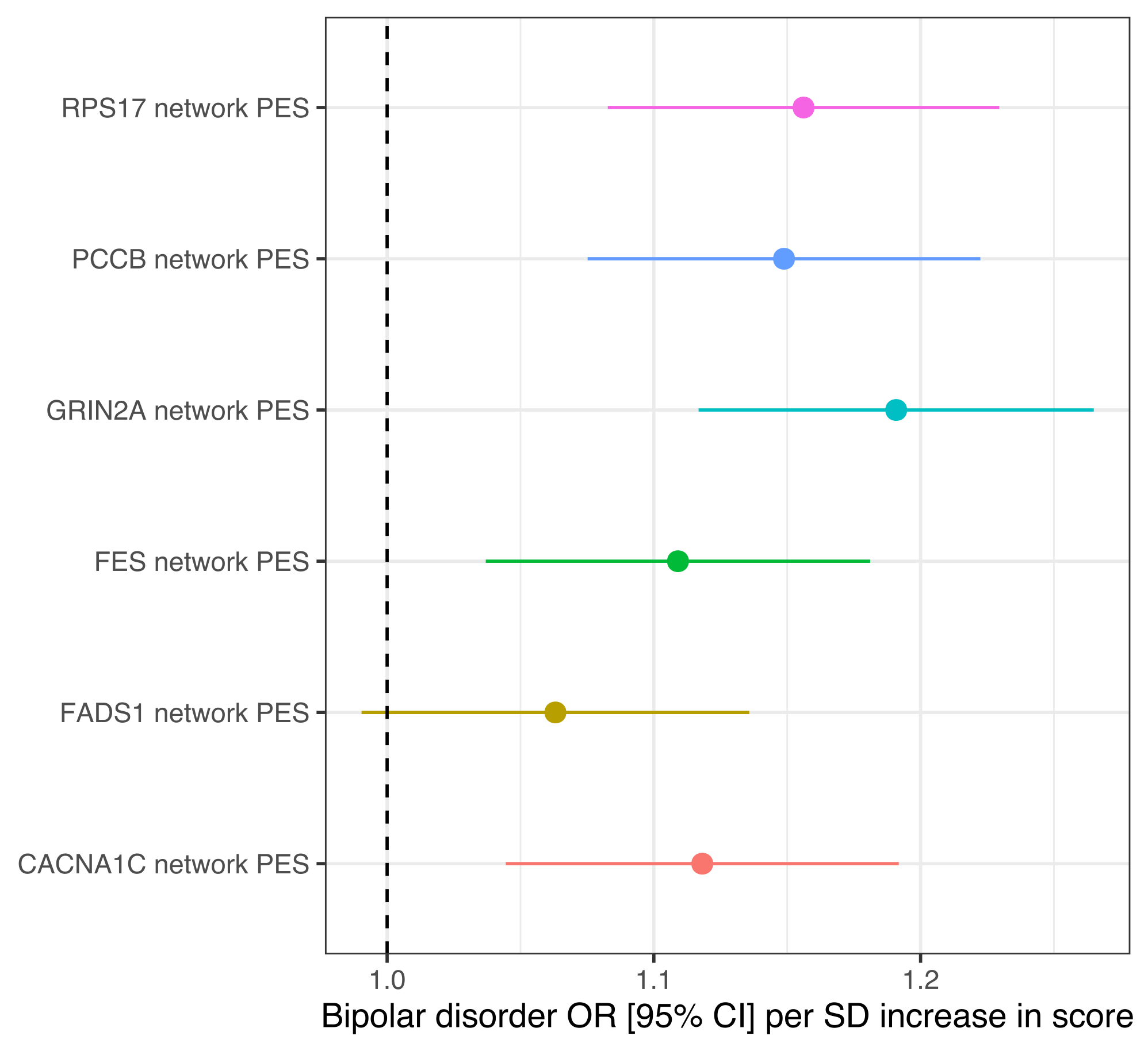

**a**

**bb**

**Supplementary Figure 1.** **Network PES** **effect sizes in the training set for schizophrenia and bipolar disorder**. Forest plot of odds ratio for schizophrenia (**a**) and bipolar disorder (**b**) per standard deviation increase in each score is presented. Error bars denote 95% confidence intervals.

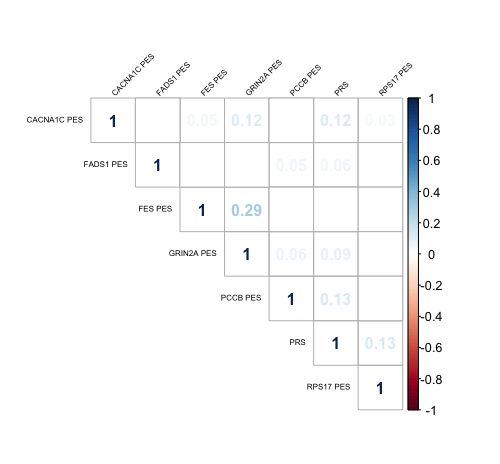

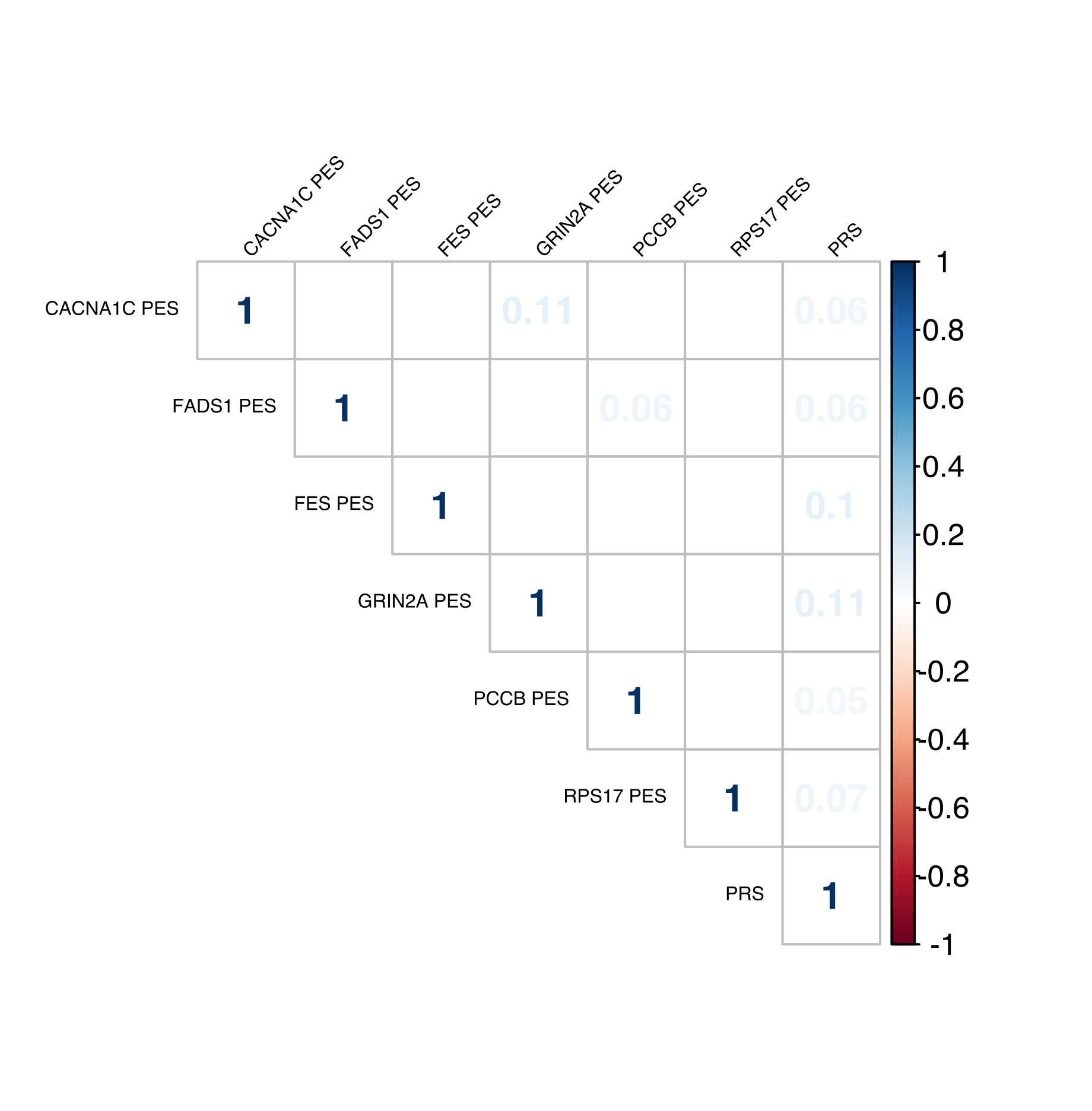

**a**

**b**

**Supplementary Figure 2**. **Correlation plot (Pearson’s) amongst network PES and genome wide PRS**. Nominally significant correlations (*P* < 0.05) have their correlation coefficients displayed. The top panel (**a**) is for schizophrenia scores and the bottom panel (**b**) is for BIP scores.
